# Night-to-night REM sleep variability: a relevant marker of early amyloid-β deposition

**DOI:** 10.64898/2025.12.19.25342684

**Authors:** Blandine Montagne, Maéva Boulin, Anaïs Hamel, Pierre Champetier, Stéphane Rehel, Florence Mézenge, Brigitte Landeau, Marion Delarue, Oriane Hébert, Célia Soussi, Françoise Bertran, Gaël Chételat, Claire André, Géraldine Rauchs, the Medit-Ageing Research Group

## Abstract

**INTRODUCTION:** Sleep disturbances are prevalent in patients with Alzheimer’s disease (AD) and may emerge before overt clinical symptoms. We characterized sleep alterations in cognitively unimpaired older adults with cerebral amyloid deposition, and assessed their associations with regional amyloid deposition, cognitive and psychoaffective outcomes.

**METHODS:** Seventy-six older adults (69.1 ± 3.4 years, 63.2% female) underwent a multi-night (4.5 ± 0.8 nights) objective sleep assessment using the Somno-Art^®^ wearable device, Florbetapir-PET scanning, and an extensive neuropsychological and psychoaffective evaluation.

**RESULTS:** Amyloid β (Aβ)-positive individuals had a shorter total sleep time (TST) and greater night-to-night variability in rapid eye movement (REM) sleep duration than Aβ-negative individuals. Across the whole sample, these sleep characteristics were associated with increased Aβ deposition in widespread brain regions, but not with cognitive or psychoaffective measures.

**DISCUSSION:** Shorter sleep duration and greater REM sleep variability may index early AD-related brain changes, warranting longitudinal studies to establish their prognostic significance.

Age-Well randomized clinical trial of the Medit-Ageing European Project. Trial registration number: EudraCT:2016–002,441–36; IDRCB:2016-A01767–44; ClinicalTrials.gov Identifier: NCT02977819

## 1 BACKGROUND

The accumulation of Alzheimer’s disease (AD) pathology, characterized by abnormal deposition of amyloid β peptide (Aβ) and neurofibrillary tangles, begins years before clinical manifestations appear. Individuals with abnormal brain Aβ deposition, even without symptoms, are considered at risk for progression to AD [1] and represent a population of interest for preventive interventions.

Sleep and circadian disturbances are highly prevalent in AD patients, and may appear years before clinical symptoms manifest [2]. These disturbances notably include reduced and more fragmented nocturnal sleep, increased sleep latency and wake after sleep onset, along with reduced sleep efficiency and slow wave sleep (SWS or N3 sleep) [2–4]. Additionally, rapid eye movement (REM) sleep shows specific alterations notably related to the dysfunction of cholinergic neurotransmission [5,6]. Importantly, growing evidence supports a bidirectional relationship between sleep and neurodegeneration. Sleep disturbances may not only result from neuronal and synaptic alterations in sleep-regulating brain regions, but also contribute to AD pathology, notably by favoring the cerebral accumulation of neurotoxic metabolites, such as the Aβ peptide. This accumulation may occur through increased production and/or decreased glymphatic clearance [2,7]. Various sleep parameters have been associated with amyloid deposition in cognitively unimpaired older individuals, including self-reported sleep difficulties, altered sleep architecture (e.g., shorter sleep duration, sleep fragmentation) and microstructure (e.g., lower slow wave activity during non-REM sleep, and REM sleep theta power) [8–11]. In this context, sleep disturbances are increasingly recognized as predictive of AD risk, pathology, and cognitive decline [2,7,12–15]. In addition, sleep disturbances in AD have been associated with behavioral and neuropsychiatric symptoms, such as depression and anxiety [16]. Notably, sleep disturbances, anxious-depressive symptoms, and Aβ may interact in a negative feedback loop from the early stages of the AD continuum, driving tau accumulation, neurodegeneration, and subsequent cognitive decline [17].

Nevertheless, several inconsistencies in the literature limit our ability to draw firm conclusions about the associations between sleep disturbances and early AD pathology, and the clinical consequences of such disturbances remain poorly understood. For example, a recent study of five large pooled cohorts failed to establish an association between standard measures of sleep architecture (e.g., sleep stage duration, sleep efficiency, slow wave activity) and the risk of incident dementia [18]. Discrepancies across studies may, at least in part, be attributed to the variety of sleep disturbances measured, the diversity of study populations in terms of disease severity, and methodological limitations. Indeed, heterogeneous assessment methods are used to characterize sleep quality and continuity. These include subjective assessments, which do not always reflect objective sleep measures; actigraphy data, which usually monitors rest/activity cycles averaged over several days but does not provide information about sleep stages; or single-night polysomnography measurements, which can be affected by setting-related factors. A recent Delphi consensus study conducted by the *Sleep and Circadian Rhythms Professional Interest Area* of the International Society to Advance Alzheimer’s Research and Treatment (ISTAART) and international experts [19] recommended prioritizing more continuous assessments of sleep and identified night-to-night sleep variability as a highly relevant aspect of sleep on which future research should focus. Indeed, greater variability in sleep patterns (e.g., sleep fragmentation) has been linked to AD risk, underlying pathology (including Aβ burden), and cognitive impairment [20–25]. Yet, no study has specifically examined the night-to-night variability in sleep stages. The emergence of ecological sleep-tracking technologies provides a unique opportunity to address this gap, by enabling multi-night, at-home recordings of sleep architecture. Such devices allow the investigation of the intra-individual variability of sleep metrics in relation to early AD pathophysiological processes.

The objectives of the current study were thus to i) identify changes in mean levels and night-to-night variability of sleep architecture in cognitively unimpaired older adults with cerebral amyloid deposition, ii) map the regional associations between these sleep metrics and amyloid burden, and iii) explore their associations with cognitive and psychoaffective outcomes.

## 2 METHODS

### 2.1 Participants

The present study was conducted using data from the baseline visit of the Age-Well randomized controlled trial (RCT) of the Medit-Ageing European project, sponsored by the French National Institute of Health and Medical Research (INSERM). The Age-Well RCT was approved by the ethics committee (CPP Nord-Ouest III, Caen; trial registration number: EudraCT: 2016-002441-36; IDRCB: 2016-A01767-44; Clinicaltrials.gov Identifier: NCT02977819). All participants provided written informed consent prior to the examinations.

Participants were enrolled between November 24, 2016, and March 5, 2018. Detailed biological, behavioral, neuroimaging, and sleep data were collected in Caen, France. The protocol and eligibility criteria have been previously described [26]. Briefly, participants were 65 years or older, community-dwelling, native French speakers, retired for at least one year, had a minimum of 7 years of formal education, and performed within the normal range for their age and educational level on standardized cognitive tests. The main exclusion criteria included safety concerns related to magnetic resonance imaging (MRI) or positron emission tomography (PET), evidence of a major neurological or psychiatric disorder (including substance use disorder), history of cerebral disease, presence of a chronic disease or acute unstable illness, and current or recent medication that may interfere with cognitive functioning. Two participants were excluded from all secondary analyses by the Trial Steering Committee for not meeting the eligibility criteria (i.e., history of head trauma and diagnosis of amyotrophic lateral sclerosis during the study [likely in a subclinical state at inclusion]).

At baseline, and within a maximum period of 3 months, participants underwent a structural MRI, a Florbetapir PET scan, Apolipoprotein E (*APOE*) genotyping, in-home polysomnography (PSG), and a sleep recording using the Somno-Art^®^ wearable device for up to 7 consecutive nights, as well as neuropsychological and psychoaffective assessments.

### 2.2 Neuroimaging Data Acquisition and Analysis

Participants underwent structural MRI and ^18^F-Florbetapir (AV45, Amyvid) PET scans at the Cyceron Center (Caen, France), using a Philips Achieva 3T Scanner and a Discovery RX VCT 64 PET-CT scanner (General Electric Healthcare), respectively. Details on data acquisition and processing have been previously described [27]. Briefly, T1-weighted anatomical images were acquired using a 3D fast-field echo sequence (3D-T1-FFE sagittal, repetition time (TR) = 7.1 ms, echo time (TE) = 3.3 ms, flip angle = 6°, 180 slices with no gap, slice thickness = 1 mm, field of view (FoV) = 256 × 256 mm^2^, in-plane resolution = 1 × 1 mm^2^), segmented using Fluid Attenuated Inversion Recovery images (3D-IR sagittal, TR = 4,800 ms, TE = 272 ms, inversion time (TI) = 1,650 ms, flip angle = 40°, 180 slices with no gap, slice thickness = 1 mm, FoV = 250 × 250 mm^2^, in-plane resolution = 0.98 × 0.98 mm^2^), spatially normalized to the Montreal Neurological Institute (MNI) template, and modulated using the SPM12 segmentation procedure.

A transmission scan was performed for attenuation correction prior to PET acquisition. A 10-minute PET scan, beginning 50 minutes after the intravenous injection of ∼4MBq/Kg of ^18^F-Florbetapir (resolution = 3.76 x 3.76 x 4.9 mm^3^, FoV = 157 mm, voxel size = 1.95 x 1.95 x 3.27 mm^3^), was then acquired to reflect amyloid deposition.

PET data were co-registered with individual T1-weighted MRI scans, voxel-wise corrected for partial volume effects (PVE) using the two-compartmental voxel-wise Müller-Gärtner method [28], and spatially normalized to the MNI template using deformation parameters derived from the T1-weighted segmentation procedure. Images were then quantitatively scaled using cerebellar gray matter as a reference, resulting in standardized uptake value ratio (SUVR) images. A smoothing kernel of 10 mm Gaussian filter was applied and images were masked to exclude non-gray matter voxels from the analyses.

Aβ status was characterized following previously described methodology [29]. Amyloid uptake was extracted and averaged across the typical regions with high amyloid load in AD, including the frontal, temporal and parietal cortices and precuneus [30]. SUVRs were then converted to the Centiloid scale following the standardized procedure of Klunk et al. [30] adapted for Florbetapir SUVRs [31]. Based on previous work [32], a threshold of 12 Centiloid was applied to dichotomize participants as Aβ-positive (Aβ+) or Aβ-negative (Aβ-).

### 2.3 Sleep Measures

Sleep was monitored at home for 3 to 7 nights (mean number of nights: 4.5 ± 0.8) using the Somno-Art^®^ wearable device (Generation 1; https://www.somno-art.com/). This armband records synchronized estimates of heart rate and movement quantification (actigraphy) during a sleep allocated period of at least 5 hours. Somno-Art^®^ Software v.2.4.0 [2.4.1] was used to analyze heart rate estimates (with beat-to-beat resolution) and actigraphy signals (with 1-Hz resolution), providing sleep stage classification at a 1-second epoch resolution. The latter classification was resampled in 30-second epochs, selecting the dominant stage (or the first stage to appear if stages were equally represented). The sleep classification algorithm is based on expert rules associated with Support Vector Machine (SVM) detectors. Further information on the data processing methodology is available in Muzet et al. [33]. The main exclusion criteria for the recordings were no or incorrect use of the device (e.g., device removal), failure to import data, time-stamping issues, invalid recordings for software analysis (i.e., bad actigraphy and/or heart rate signals), exclusion from quality control (e.g., time in bed less than 5 hours), or less than 3 valid recordings available for one individual. Somno-Art^®^ Software has been validated against PSG visual scoring in healthy young to middle-aged adults and patients with insomnia, obstructive sleep apnea, and major depressive disorder [33,34]. We evaluated the performance of Somno-Art^®^ Software in an elderly population, by conducting comparison analyses between Somno-Art^®^ Software classification and PSG visual scoring in a subsample of the Age-Well RCT with matching Somno-Art^®^ and PSG recordings. The detailed methodology and outcomes are available in Supplementary Methods. Briefly, the discrepancy analysis revealed a proportional bias for all parameters, the magnitude of the bias depending on the PSG-derived measures (Table S1). Bland-Altman plots are displayed in Figure S1. Regarding Wake/Sleep classification, the epoch-by-epoch analysis revealed that Somno-Art^®^ Software exhibited 91% sensitivity (i.e., ability to correctly classify PSG sleep epochs) and 54% specificity (i.e., ability to correctly classify PSG wake epochs), with a “substantial” agreement between measurements (prevalence-adjusted bias-adjusted kappa (PABAK): 0.64). For the four-stage classification, agreement between the two methods ranged from “fair” for N1+N2 (PABAK: 0.28) to “substantial” for Wake (PABAK: 0.64), N3 (PABAK: 0.72), and REM (PABAK: 0.68) stages. A proportional error matrix between the two methods is presented in Table S2 and additional epoch-by-epoch metrics are provided in Table S3.

Several metrics reflecting sleep architecture and continuity were derived from the sleep stage classification: total sleep time (TST, min; corresponding to the total duration of non-wake epochs), sleep onset latency (SOL, min; corresponding to the time from lights-off to the first sleep epoch), wake after sleep onset (WASO, min; corresponding to the duration of wake epochs after sleep onset), sleep efficiency (SE, %; corresponding to the ratio between total sleep time and time in bed), the duration of N1, N2, N3 and REM sleep stages (expressed in minutes and as a percentage of TST), and REM sleep latency (REML, min; corresponding to the duration between sleep onset and the first epoch of REM sleep). For each parameter, mean and night-to-night variability measures were calculated. The intra-individual variability (IIV) of sleep parameters was characterized using an intra-individual coefficient of variation, computed as the standard deviation divided by the mean, multiplied by 100. This mean-referenced measure of variability accounts for the greater likelihood of higher variability in individuals with higher mean scores [23,35–37].

Additionally, self-reported sleep quality and disturbances, as well as insomnia symptoms, were assessed using the Pittsburgh Sleep Quality Index (PSQI) [38] and the Insomnia Severity Index (ISI) [39], respectively.

### 2.4 Cognitive and Psychoaffective Measures

All participants underwent a detailed neuropsychological assessment, providing a number of scores reflecting global functioning, episodic and working memory and various executive functions such as inhibition, flexibility, and processing speed. We used those scores to create four cognitive composite scores targeting cognitive domains typically affected in aging and early AD: the Preclinical Alzheimer’s Cognitive Composite 5 (PACC-5), verbal episodic memory, executive functioning, and attention/speed (see Supplementary Methods, [26]). Additionally, subjective cognitive difficulties were evaluated with the Cognitive Difficulties Scale (CDS) [40]. Two subscores were obtained from the CDS: 1) a reduced score reflecting global cognitive difficulties, and 2) a score specifically reflecting memory difficulties, with higher values indicating greater perceived difficulties (details in Supplementary Methods).

Psychoaffective measures included the 15-item version of the Geriatric Depression Scale (GDS) [41] to highlight subclinical depressive symptoms, and the Form Y-B of the State-Trait Anxiety Inventory (STAI-B) [42] to assess trait anxiety symptoms. In addition, two complementary aspects of repetitive negative thinking were measured: 1) brooding (i.e., past-directed negative thoughts) and 2) worry (i.e., future-directed negative thoughts), respectively assessed using the Brooding subscore of the Rumination Response Scale (RRS) [43] and the total score of the Penn State Worry Questionnaire (PSWQ) [44]. Higher scores on these psychoaffective questionnaires indicate greater severity of symptoms (details in Supplementary Methods).

### 2.5 Statistical Analyses

Statistical analyses were performed using R (version 4.4.2; http://www.r-project.org) and SPM12. Differences between groups for demographics and subjective sleep measures according to Aβ status were assessed using non-parametric Wilcoxon rank-sum test for continuous variables and chi-square statistics (or a non-parametric Fisher’s exact test) for categorical variables. For all analyses, statistical significance was set to *P* < .05, except for voxel-wise multiple regressions. For the latter, results were considered significant at a *P* < .005 (uncorrected) threshold, with a minimal cluster size of k = 100 voxels.

To identify sleep characteristics altered in cognitively unimpaired older individuals with cerebral amyloid deposition, all sleep parameters were compared between Aβ- and Aβ+ participants using type II analyses of covariance (ANCOVAs), with Aβ status as the between-participants factor and sleep metrics as the dependent variables. For models that did not meet the conditions for ANCOVA, robust ANCOVA models were additionally performed using *lmrob* function.

Then, to characterize the cerebral topography of the associations between these Aβ-related sleep metrics and Aβ deposition, voxel-wise multiple regression analyses were performed between each Aβ-related sleep metric and amyloid load obtained from PET images. Lastly, we assessed the associations between Aβ-related sleep metrics and cognitive and psychoaffective outcomes by conducting multiple regression analyses with cognitive and psychoaffective measures as dependent variables and sleep metrics as independent predictors. Robust linear regressions were performed to confirm the results of models that did not meet the conditions for multiple linear regression.

All analyses were adjusted for age, sex, education level, and the apnea-hypopnea index (AHI) obtained from a polysomnography recording performed as part of the protocol. For significant results, secondary sensitivity analyses were performed by additionally controlling for *APOE* ε4 status (i.e., carrying at least one copy of the ε4 allele) and the number of recorded nights.

## 3 RESULTS

### 3.1 Sample characteristics

Of the 135 participants enrolled in the Age-Well RCT at baseline, 76 underwent sleep assessments and neuroimaging scans, neuropsychological and psychoaffective examinations, as well as blood sampling for *APOE* ε4 genotyping, and were included in the final analysis sample (flow diagram in Figure 1). Their characteristics are described in Table 1. The study sample consisted of cognitively unimpaired older adults with an average age of 69.1 ± 3.4 years, 63.2% female, 22.4% *APOE* ε4 carriers, and an average education level of 12.8 ± 3.0 years. Fifty-four individuals were Aβ-, and 22 were Aβ+. There were no significant differences between the two groups in terms of age, sex ratio, education level, Mini-Mental State Examination (MMSE) score, body mass index (BMI), number of recorded nights, proportion of *APOE* ε4 carriers, or current use of sleep medication (i.e., on a regular basis (≥1/week), as determined by the PSQI item about sleep medication use during the last month). As expected, the Aβ+ group displayed a greater Centiloid value (W = 1,188; *P* < .001). Additionally, Aβ+ participants exhibited higher AHI values (W = 843.5; *P* = .004). Interestingly, no differences were observed between groups in terms of subjective sleep quality or the severity of insomnia symptoms, as assessed by the PSQI and the ISI, respectively. The descriptive characteristics of participants’ sleep metrics are presented in Table 2, and a correlation matrix between all sleep metrics (mean and variability) is provided in Figure S2.

**FIGURE 1.**
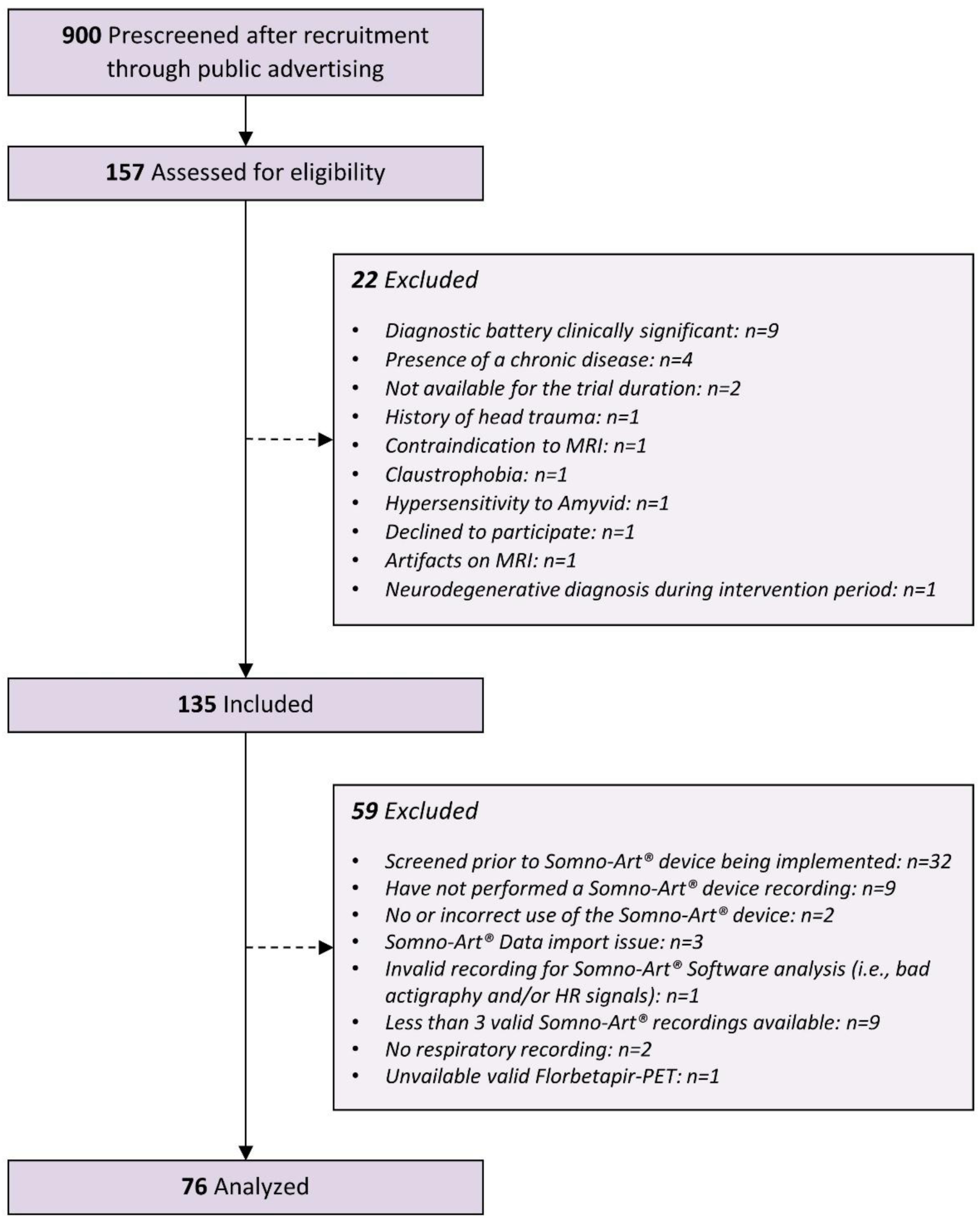
Flow diagram of the inclusion process. Abbreviations: HR, heart rate; MRI, magnetic resonance imaging; PET, positron emission tomography.

**TABLE 1.** Participants’ characteristics.

| Characteristic | Total sample (n = 76) | Aβ- (n = 54) | Aβ+ (n = 22) | *P* value for between-group differences |
| --- | --- | --- | --- | --- |
| Age (years) | 69.1 ± 3.4 [65.0; 79.0] | 68.8 ± 3.2 [65.0; 77.6] | 69.6 ± 4.0 [65.0; 79.0] | .499 |
| Women, n (%) | 48 (63.2) | 34 (62.9) | 14 (63.6) | .956 |
| Education (years) | 12.8 ± 3.0 [7; 20] | 12.9 ± 3.0 [7; 17] | 12.7 ± 3.2 [7; 20] | .758 |
| MMSE total score | 29.0 ± 1.1 [26; 30] | 29.1 ± 1.0 [26; 30] | 28.9 ± 1.1 [27; 30] | .391 |
| BMI (kg/m<sup>2</sup>) | 25.7 ± 3.9 [18.1; 37.2] | 25.6 ± 3.8 [18.1; 37.2] | 26.1 ± 4.1 [20.4; 33.9] | .762 |
| Current sleep medication use, n (%) <sup>a</sup> | 3 (4.0) | 3 (5.6) | 0 (0.0) | .552 |
| AHI (n/h) | 25.3 ± 14.1 [2.4; 75.5] | 22.0 ± 11.6 [2.4; 60.5] | 33.5 ± 16.9 [10.4; 75.5] | **.004** |
| *APOE* ε4 carrier, n (%) | 17 (22.4) | 10 (18.5) | 7 (31.8) | .234 |
| Centiloid value | 9.0 ± 26.0 [-22.0; 101.3] | -3.6 ± 8.8 [-22.0; 11.8] | 40.1 ± 28.1 [12.1; 101.3] | **<.001** |
| PSQI total score | 4.9 ± 3.3 [0; 12] | 4.8 ± 3.4 [0; 12] | 5.0 ± 3.2 [1; 12] | .652 |
| ISI total score | 6.9 ± 5.2 [0; 20] | 6.4 ± 4.8 [0; 19] | 8.1 ± 6.0 [1; 20] | .307 |
| Number of Somno-Art<sup>®</sup> recorded nights | 4.5 ± 0.8 [3; 7] | 4.6 ± 0.7 [3; 6] | 4.3 ± 1.1 [3; 7] | .285 |

NOTE. Data are presented as mean ± SD [minimum; maximum] for continuous variables or n (%) for categorical variables. Differences between groups for demographics and subjective sleep measures according to Aβ status were assessed using non-parametric Wilcoxon rank-sum test for continuous variables and chi-square statistics (or a non-parametric Fisher's exact test) for categorical variables. Results in bold indicate significant differences at the *P* < .05 level.
<sup>a</sup> Use of sleep medication on a regular basis ( $\geq 1$ /week), as determined by the PSQI item about sleep medication use during the last month.
Abbreviations: %, percentage; Aβ , amyloid $\beta$ ; AHI, apnea–hypopnea index; *APOE* ε4, ε4 allele of the Apolipoprotein E; BMI, body mass index; h, hour; ISI, Insomnia Severity Index; MMSE, Mini-Mental State Examination; n, number; PSQI, Pittsburgh Sleep Quality Index; SD, standard deviation.

**TABLE 2.** Descriptive characteristics of sleep mean and variability metrics.

| Sleep parameter | Mean value | Intra-individual variability (IIV, %) |
| --- | --- | --- |
| TST (min) | 395.0 ± 54.0 [231.7; 501.3] | 11.4 ± 5.3 [3.0; 30.1] |
| SOL (min) | 19.4 ± 12.4 [1.2; 75.7] | 68.3 ± 29.6 [15.8; 153.8] |
| WASO (min) | 69.9 ± 37.7 [23.4; 227.0] | 36.8 ± 18.1 [3.0; 99.8] |
| SE (%) | 81.7 ± 8.0 [48.6; 92.1] | 7.0 ± 4.1 [1.7; 22.2] |
| N1 (min) | 35.2 ± 8.3 [15.7; 64.8] | 18.6 ± 7.5 [2.8; 40.2] |
| N1 (%) | 9.2 ± 2.7 [4.0; 17.3] | 18.6 ± 7.4 [5.4; 47.4] |
| N2 (min) | 216.6 ± 35.6 [114.8; 320.7] | 16.1 ± 7.7 [3.3; 40.1] |
| N2 (%) | 54.9 ± 6.0 [41.4; 68.6] | 10.9 ± 5.1 [3.5; 31.6] |
| N3 (min) | 68.2 ± 30.2 [4.7; 162.1] | 37.7 ± 19.2 [4.8; 89.3] |
| N3 (%) | 17.1 ± 6.7 [1.5; 33.7] | 36.1 ± 17.9 [11.0; 83.2] |
| REM (min) | 75.0 ± 18.6 [33.9; 115.1] | 23.6 ± 11.5 [5.5; 56.7] |
| REM (%) | 18.9 ± 3.4 [10.9; 25.1] | 20.5 ± 10.1 [2.7; 48.5] |
| REML (min) | 86.5 ± 29.5 [43.2; 174.0] | 41.4 ± 20.4 [8.4; 88.4] |

NOTE. Data are presented as mean ± SD [minimum; maximum]. Intra-individual coefficients of variation are computed as the standard deviation of the sleep metric divided by the mean, multiplied by 100.
Abbreviations: %, percentage; IIV, intra-individual variability; min, minutes; REM, rapid eye movement sleep; REML, rapid eye movement sleep latency; SD, standard deviation; SE, sleep efficiency; SOL, sleep onset latency; TST, total sleep time; WASO, wake after sleep onset.

### 3.2 Differences in sleep architecture according to Aβ status

When comparing sleep parameters between the two groups (Aβ+ vs Aβ-) using ANCOVAs (Table 3), adjusting for age, sex, education and the AHI, Aβ+ individuals exhibited shorter TST (F = 4.85, *P* = .031, η_p_^2^ = 0.06 and greater night-to-night variability of REM sleep duration (REM-IIV, min: F = 5.31, *P* = .024, η_p_^2^ = 0.07; REM-IIV, %: F = 4.07, *P* = .048, η_p_ ^2^ = 0.05). Results remained unchanged with robust ANCOVAs for models that did not meet criteria for ANCOVA (Table S4). No other significant differences were observed.

**TABLE 3.** Comparisons of sleep mean and variability metrics between Aβ- and Aβ+ individuals.

| Sleep parameter | Df | Aβ- adjusted mean [95% CI] | Aβ+ adjusted mean [95% CI] | F | *P* | $\eta_p^2$ |
| --- | --- | --- | --- | --- | --- | --- |
| ***Mean Sleep Metrics*** |  |  |  |  |  |  |
| TST (min) | 1 | 397.1 [383.8, 410.4] | 369.0 [347.8, 390.2] | 4.85 | **.031** | 0.065 |
| SOL (min) | 1 | 20.5 [17.0, 24.1] | 17.8 [12.2, 23.5] | 0.65 | .423 | 0.009 |
| WASO (min) | 1 | 72.5 [62.5, 82.5] | 68.8 [52.7, 84.9] | 0.15 | .700 | 0.002 |
| SE (%) | 1 | 81.1 [78.9, 83.2] | 81.1 [77.7, 84.5] | 0.00 | 1.00 | $6.414 \times 10^{-10}$ |
| N1 (min) | 1 | 36.1 [33.9, 38.4] | 34.0 [30.4, 37.5] | 1.01 | .318 | 0.014 |
| N1 (%) | 1 | 9.4 [8.7, 10.1] | 9.5 [8.5, 10.6] | 0.04 | .834 | $6.309 \times 10^{-4}$ |
| N2 (min) | 1 | 218.7 [209.0, 228.5] | 201.8 [186.2, 217.4] | 3.27 | .075 | 0.045 |
| N2 (%) | 1 | 55.3 [53.6, 57.0] | 54.4 [51.7, 57.1] | 0.28 | .596 | 0.004 |
| N3 (min) | 1 | 67.0 [59.1, 75.0] | 63.6 [50.9, 76.4] | 0.20 | .659 | 0.003 |
| N3 (%) | 1 | 16.5 [14.7, 18.4] | 17.4 [14.5, 20.3] | 0.24 | .628 | 0.003 |
| REM (min) | 1 | 75.2 [70.3, 80.2] | 69.6 [61.8, 77.5] | 1.42 | .237 | 0.020 |
| REM (%) | 1 | 18.8 [17.8, 19.7] | 18.7 [17.2, 20.2] | 0.02 | .892 | $2.641 \times 10^{-4}$ |
| REML (min) | 1 | 87.4 [78.9, 95.9] | 81.4 [67.8, 95.0] | 0.54 | .466 | 0.008 |
| ***Intra-Individual Variability (IIV, %) Sleep Metrics*** |  |  |  |  |  |  |
| TST (min) | 1 | 11.2 [9.8, 12.6] | 13.2 [10.9, 15.5] | 2.15 | .147 | 0.030 |
| SOL (min) | 1 | 66.5 [58.6, 74.4] | 68.9 [56.3, 81.5] | 0.10 | .750 | 0.001 |
| WASO (min) | 1 | 36.4 [31.5, 41.4] | 32.9 [24.9, 40.8] | 0.57 | .454 | 0.008 |
| SE (%) | 1 | 7.5 [6.3, 8.6] | 6.5 [4.6, 8.3] | 0.76 | .387 | 0.011 |
| N1 (min) | 1 | 19.0 [16.9, 21.1] | 16.5 [13.1, 19.9] | 1.46 | .232 | 0.020 |
| N1 (%) | 1 | 19.7 [17.6, 21.7] | 15.9 [12.5, 19.2] | 3.66 | .060 | 0.050 |
| N2 (min) | 1 | 15.7 [13.6, 17.9] | 18.6 [15.2, 22.1] | 2.01 | .161 | 0.028 |
| N2 (%) | 1 | 10.6 [9.1, 12.0] | 11.8 [9.4, 14.1] | 0.74 | .394 | 0.010 |
| N3 (min) | 1 | 39.2 [33.9, 44.5] | 36.2 [27.8, 44.7] | 0.35 | .557 | 0.005 |
| N3 (%) | 1 | 37.4 [32.5, 42.3] | 35.3 [27.4, 43.1] | 0.21 | .649 | 0.003 |
| REM (min) | 1 | 22.2 [19.2, 25.3] | 28.9 [24.1, 33.7] | 5.31 | **.024** | 0.070 |
| REM (%) | 1 | 18.9 [16.1, 21.7] | 24.3 [19.8, 28.7] | 4.07 | **.048** | 0.055 |
| REML (min) | 1 | 40.5 [34.6, 46.5] | 43.3 [33.8, 52.9] | 0.24 | .629 | 0.003 |

NOTE. Results of type II ANCOVAs performed with each sleep parameter as the dependent variable, Aβ status as the between-participants factor, and age, sex, education, and AHI as covariates. Results in bold indicate significant differences at the $P < .05$ level.
Abbreviations: %, percentage; Aβ , amyloid $\beta$ ; AHI, apnea–hypopnea index; CI, confidence interval; Df, degrees of freedom; $\eta_p^2$ , partial eta squared; IIV, intra-individual variability; min, minutes; REM, rapid eye movement sleep; REML, rapid eye movement sleep latency; SE, sleep efficiency; SOL, sleep onset latency; TST, total sleep time; WASO, wake after sleep onset.

Sensitivity analyses (Tables S5 and S6) showed similar results for TST and the variability of REM sleep duration expressed in minutes (REM-IIV, min), when additionally controlling for *APOE* ε4 status (TST: F = 5.91, *P* = .018, η_p_^2^ = 0.08; REM-IIV, min: F = 5.77, *P* = .019, η_p_^2^ = 0.08) and the number of recorded nights (TST: F = 4.47, *P* = .038, η_p_^2^ = 0.06; REM-IIV, min: F = 4.51, *P* = .037, η_p_^2^ = 0.06). However, greater variability of REM sleep duration, expressed as a percentage of TST (REM-IIV, %), observed in Aβ+ individuals, was no longer significant after additionally controlling either for *APOE* ε4 status (F = 3.96, *P* = .050, η_p_^2^ = 0.05) or the number of recorded nights (F = 3.04, *P* = .085, η_p_^2^ = 0.04). Results of sensitivity analyses remained unchanged with robust ANCOVA for the model that did not meet criteria for ANCOVA (Table S7).

### 3.3 Topography of the associations between Aβ-related sleep metrics and cerebral Aβ deposition

We then assessed the associations between sleep metrics found to be significantly altered in Aβ+ individuals (i.e., TST and the variability of REM sleep duration) and the regional topography of Aβ deposition using voxel-wise multiple regressions, adjusting for age, sex, education and the AHI. The results of significant voxel-wise regressions are presented in Figure 2, and detailed peak statistics and coordinates of significant clusters are reported in Table 4.

**FIGURE 2.**
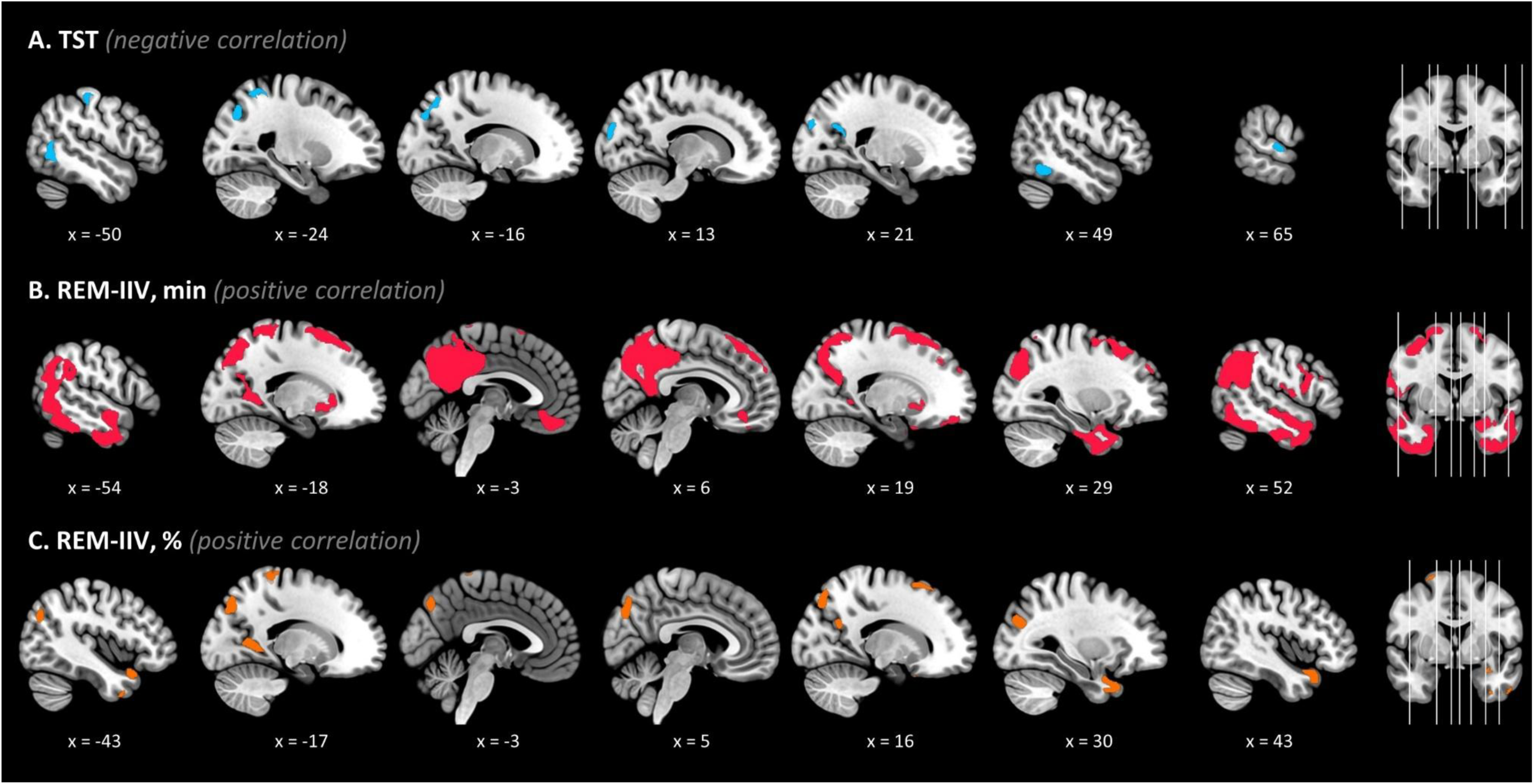
Neuroimaging patterns of significant associations between Aβ-related sleep metrics and amyloid deposition. Results of voxel-wise multiple regressions showing significant associations between (A) TST, variability in REM sleep duration expressed in (B) minutes (REM-IIV, min), and (C) as a percentage of TST (REM-IIV, %) and amyloid deposition. Results are presented at the P < .005 (uncorrected) threshold, combined with a minimal cluster size of k = 100 voxels, controlling for age, sex, education, and AHI. Abbreviations: %, percentage; Aβ, amyloid β; AHI, apnea-hypopnea index; IIV, intra-individual variability; min, minutes; REM, rapid eye movement sleep; TST, total sleep time.

**TABLE 4.**
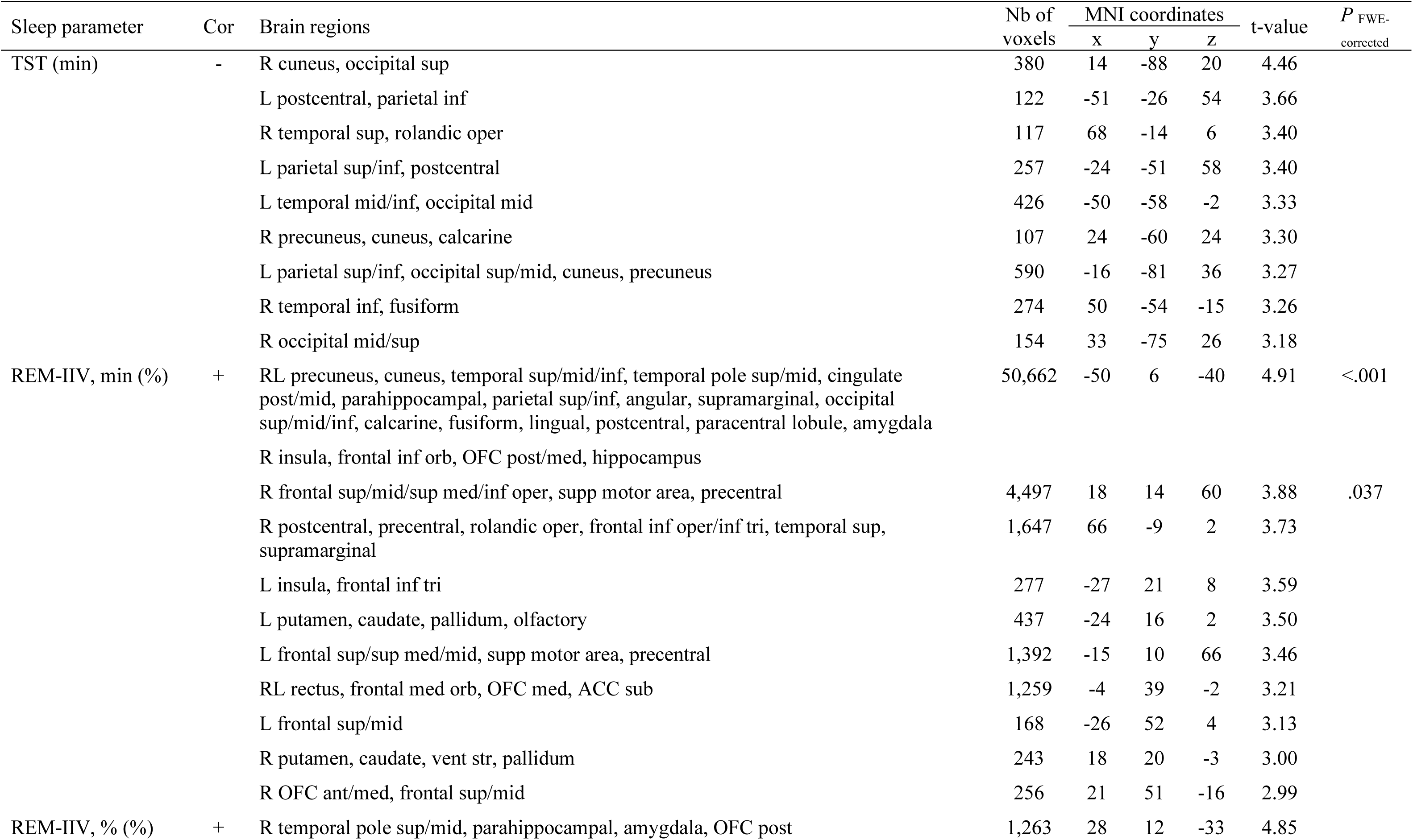
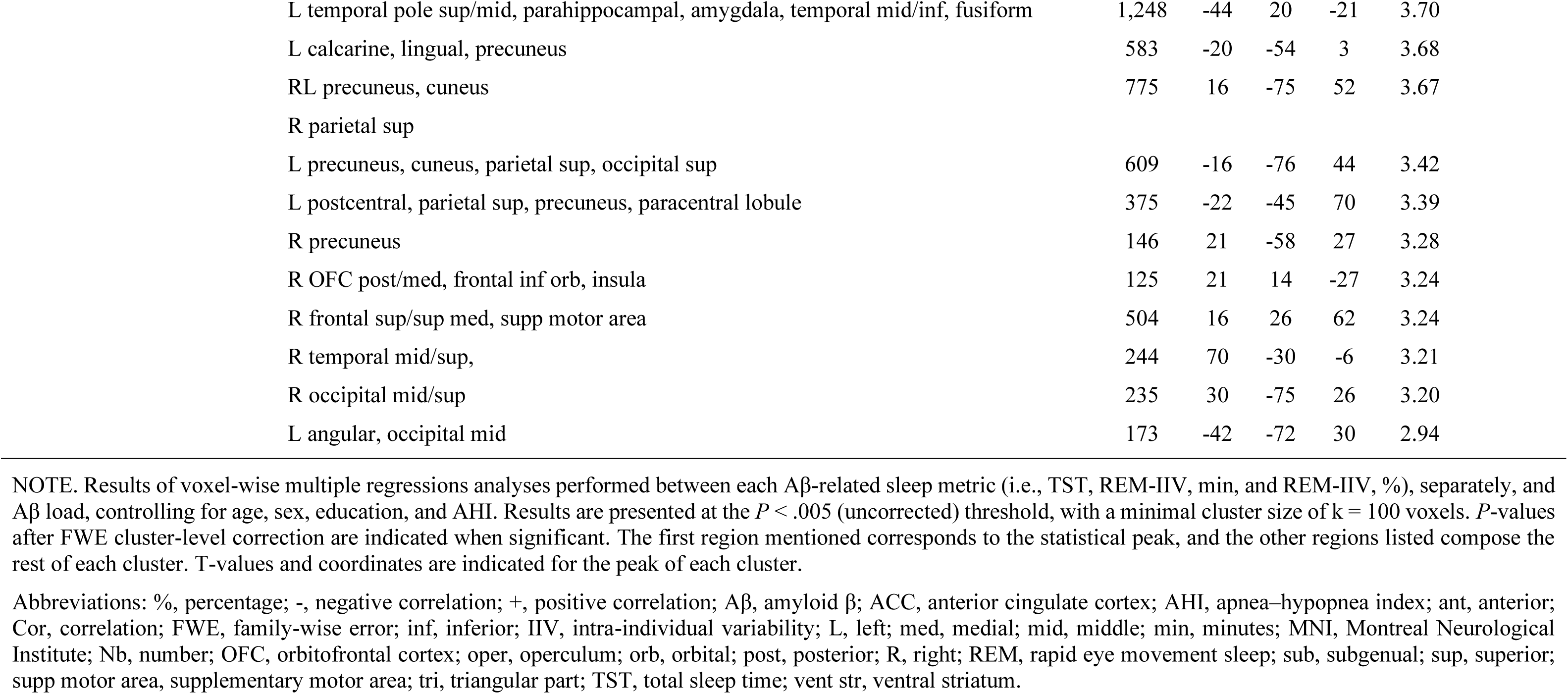
Significant associations between Aβ-related sleep metrics and amyloid deposition.

TST was negatively associated with Aβ deposition across several medial and lateral parietal and occipital areas including the cuneus and precuneus, and lateral temporal cortex. The variability of REM sleep duration, expressed in minutes (REM-IIV, min), was positively associated with Aβ deposition in widespread brain regions. A large significant cluster notably encompassed bilateral temporal and posterior regions, including the precuneus, cuneus, and posterior cingulate. Additional significant clusters were found in frontal and parietal areas, as well as subcortical regions (e.g., amygdala, pallidum). Of note, the associations observed in the main cluster and in the right frontal and precentral regions, survived a Family-Wise Error (FWE) cluster-level correction. When REM sleep duration was expressed as a percentage of TST (REM-IIV, %), its variability was associated with Aβ deposition in smaller clusters, mostly medial and lateral temporal areas and the precuneus, largely corresponding to subregions of the widespread associations observed with variability of REM sleep duration expressed in minutes (REM-IIV, min).

Overall, sensitivity analyses showed broadly consistent results after additional adjustment for *APOE* ε4 status (Table S8). After further adjusting for the number of recorded nights, the associations between TST and Aβ burden remained largely unchanged. Associations with REM sleep variability expressed in minutes (REM-IIV, min) were observed in the same regions, though to a lesser extent, and some of the associations with REM sleep variability expressed as a percentage of TST (REM-IIV, %) were no longer significant (Table S9).

### 3.4 Associations between Aβ-related sleep metrics and cognitive and psychoaffective outcomes

Lastly, multiple regression analyses were used to explore the associations between each Aβ-related sleep metric and cognitive and psychoaffective outcomes (Table 5), adjusting for age, sex, education and the AHI. No association was found between TST or variability of REM sleep duration and cognitive composite scores. We observed a marginal association between longer TST and higher subjective memory difficulties (β = 0.02; *P* = .046; Figure 3). Finally, no significant association was found between Aβ-related sleep metrics and psychoaffective outcomes. None of these results were modified using robust regressions for models that did not satisfy the conditions for regression (Table S10).

**FIGURE 3.**
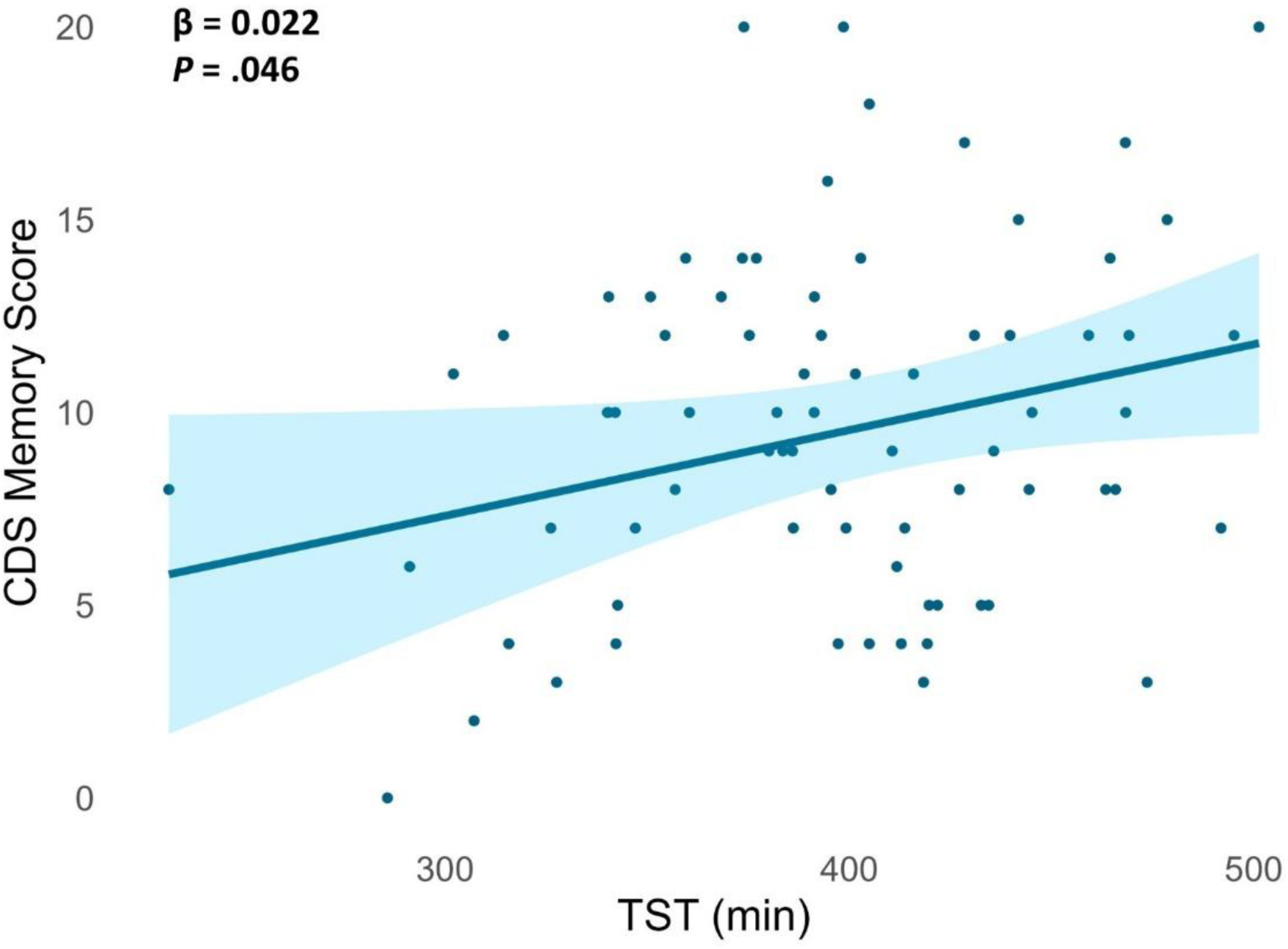
Significant association between total sleep time and subjective memory difficulties. Scatterplot illustrating the association between TST and CDS memory score, corrected for age, sex, education, and AHI. Abbreviations: AHI, apnea–hypopnea index; CDS, Cognitive Difficulties Scale; min, minutes; TST, total sleep time.

**TABLE 5.**
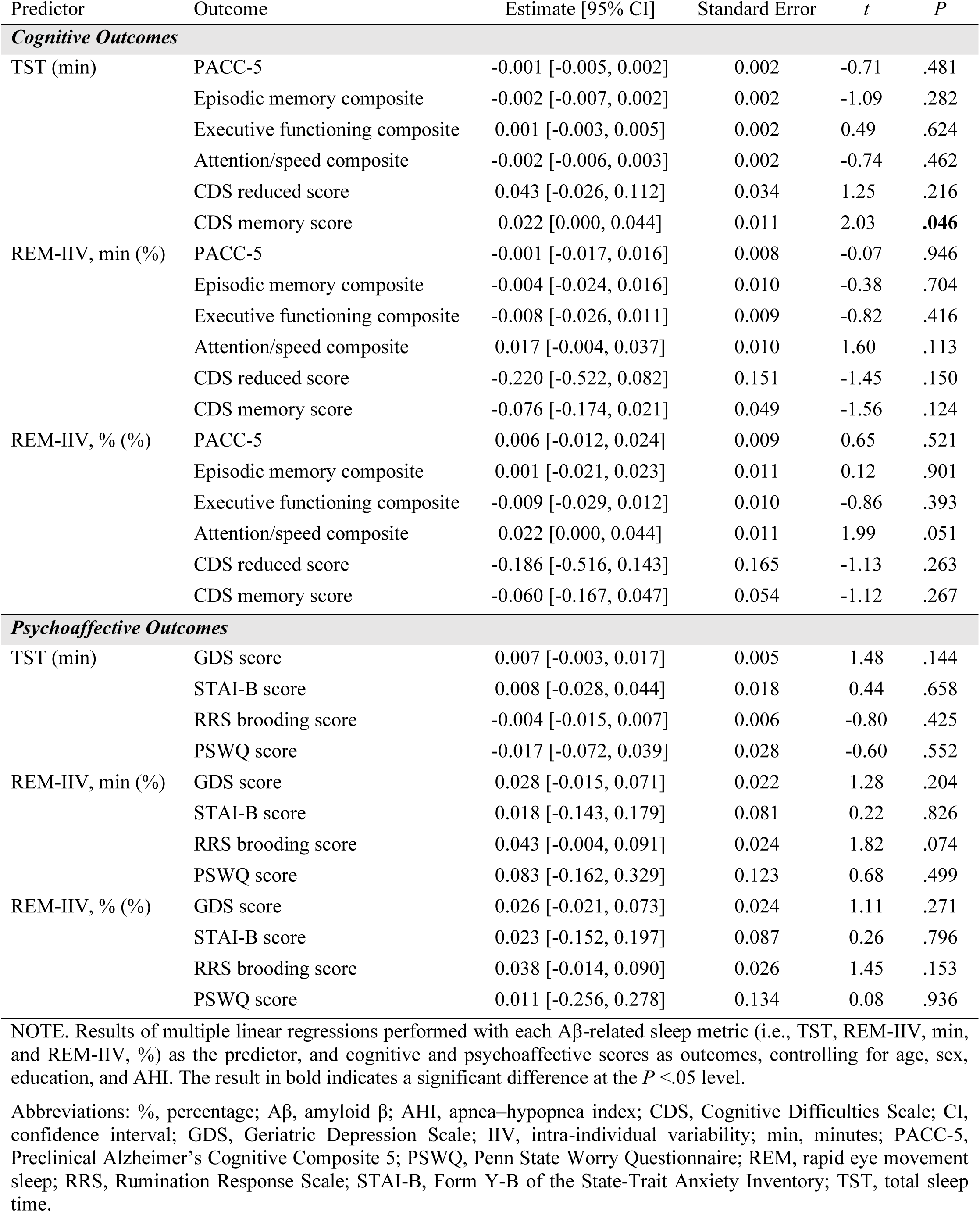
Associations between Aβ-related sleep metrics and cognitive and psychoaffective outcomes.

Sensitivity analyses showed that the association between TST and subjective memory difficulties remained significant after additional adjustment for *APOE* ε4 status (β = 0.03; *P* = .026; Table S11) or the number of recorded nights (β = 0.02; *P* = .032; Table S12).

## 4 DISCUSSION

This study aimed to characterize alterations in sleep architecture in cognitively unimpaired older adults according to Aβ status, using an ecological multi-night objective sleep assessment, and to examine their associations with regional amyloid deposition, cognitive and psychoaffective outcomes. Aβ+ individuals showed reduced mean TST and greater night-to-night variability of REM sleep duration, associated with amyloid deposition mainly in temporal and posterior regions, but not with objective cognitive or psychoaffective measures.

Our results corroborate previous studies showing that shorter sleep duration is linked to increased Aβ deposition [45–47]. This aligns with the hypothesis that sleep promotes Aβ clearance, while insufficient sleep may disrupt this process, contributing to Aβ accumulation [2]. Interestingly, shorter TST has also been associated with a higher risk of dementia [48,49]. However, a meta-analysis found no overall association between TST and amyloid load [50]. Heterogeneity in study design, sleep measurement, biomarker assessment, and statistical approaches may account for such discrepancies.

Beyond standard sleep metrics, our study identified greater night-to-night variability in REM sleep duration as a distinctive feature of Aβ+ individuals. Night-to-night variability is an increasingly recognized dimension of sleep health, with emerging evidence linking greater variability in several sleep metrics, such as sleep fragmentation, to amyloid deposition [20,23], plasma and cerebrospinal fluid (CSF) AD biomarkers [21], as well as cognitive impairment [23], dementia risk [25], and mortality [51]. Previous work investigating sleep variability has focused on sleep continuity metrics, such as sleep fragmentation, WASO or TST. For example, variability in sleep fragmentation and WASO during the first half of the night has been associated with amyloid deposition notably in frontal and parietal regions [20,24], whereas variability in TST has been associated with cerebral Aβ burden or CSF and plasma p-tau/Aβ ratios in individuals with mild cognitive impairment (MCI) [23] or a parental history of sporadic AD [21], respectively. In line with Jouvencel et al. [24], we did not observe differences in WASO or TST variability according to Aβ status, suggesting that greater variability in sleep continuity may occur later or under specific genetic or disease vulnerability conditions. Moreover, we did not identify variability in SE as an Aβ-related sleep metric in cognitively unimpaired older individuals, contrasting with prior work showing an association between SE variability and Aβ burden in MCI, especially in *APOE* ε4 carriers [23]. Of note, these results were not confirmed in individuals with a parental history of sporadic AD [21]. These data suggest that distinct variability metrics, reflecting unstable sleep patterns, may be differentially related to amyloid pathology depending on disease stage or genetic susceptibility.

REM sleep is known to play a crucial role in memory processing, emotional regulation, neural homeostasis, and synaptic regulation [52]. Previous work suggests that reduced REM sleep duration [53–55], longer REM sleep latency [54,56], or alterations in REM sleep microstructure [8], may represent early markers of AD pathology, even before dementia onset, and/or be predictive of cognitive decline and dementia risk. The mechanisms underlying REM sleep alterations likely involve early neurodegeneration within REM-regulatory networks, as core REM sleep regulatory centers (i.e., brainstem and basal forebrain cholinergic nuclei, hypothalamus) are among the earliest regions to accumulate AD-related pathology [57–61]. Recent studies suggest that resulting cholinergic denervation and neurotransmitter imbalance may compromise REM sleep integrity and stability [5,6,62].

Moreover, circadian dysfunction observed in the early stages of AD [63] may also contribute to the night-to-night instability in REM sleep duration. The daily expression of REM sleep is modulated by circadian rhythms, and may involve both promoting and inhibitory mechanisms mediated by the suprachiasmatic nucleus and orexinergic neurons [64]. Thus, AD pathology accumulation in circadian regulating areas (e.g., suprachiasmatic nucleus in the hypothalamus) [2] may lead to irregular REM propensity and fragmentation, and inconsistent REM sleep patterns across nights. Consistently, a reduction in the circadian amplitude of REM sleep has been associated with altered microstructural integrity of the hypothalamus [65]. Additionally, a dysregulation of the orexinergic system, characterized by higher CSF-orexin levels, has also been linked to REM sleep reduction in MCI patients [66]. Furthermore, our results highlight that REM sleep instability is related to amyloid accumulation in the precuneus, posterior cingulate, orbitofrontal cortex, and the insula, which are among the first to exhibit amyloid deposition in AD [67]. This finding complements our previous observations that changes in REM sleep microstructure were associated with amyloid deposition in frontal, cingulate and posterior regions in a partially overlapping sample [8]. Taken together, these results point toward REM sleep impairment as a marker of pathological aging. Of note, one might have expected SWS to be altered in Aβ+ individuals, as SWS alterations are thought to impair Aβ clearance [68] and have been associated with early amyloid deposition [11,69]. In our study, however, we did not observe such associations, likely because the wearable device only provided information on sleep architecture and did not allow us to quantify microstructural features such as slow-wave activity. Our findings of REM sleep instability should therefore be viewed as complementary to this literature: both REM sleep disruption and alterations in slow-wave activity may represent early markers of AD pathology, a possibility that needs to be clarified in future studies combining macro- and microstructural sleep measures.

Furthermore, neither TST nor REM sleep variability were associated with objective cognitive performance. This may reflect the fact that our sample comprised relatively highly educated older adults without cognitive deficits. Moreover, previous studies linking extreme sleep durations to cognitive impairment typically consider TST as a categorical factor, with both insufficient and excessive sleep being associated with impaired cognitive function [70]. Here, no participant showed abnormally long sleep duration (>9h), such that we analyzed TST as a continuous variable. Still, we found that longer TST was associated with greater subjective memory difficulties. As longer TST has been previously associated with dementia risk [71], we cannot exclude that the observed association with greater subjective memory difficulties may reflect early signs of cognitive impairment, not yet objectified. Replication and longitudinal analyses are necessary to confirm this hypothesis. Of note, REM sleep alterations and irregular sleep patterns have also been associated with increased risk of cognitive decline and/or dementia [12,25]. Here, we did not observe any association between variability in REM sleep duration and cognitive performances. Tasks specifically designed to assess cognitive functions dependent on REM sleep, such as emotional memory processing, may better capture subtle cognitive changes associated with early REM sleep alterations.

In addition, we found no association between Aβ-related sleep metrics and psychoaffective outcomes (i.e., anxiety, depressive symptoms and repetitive negative thinking), despite the fact that REM sleep is known to be involved in emotion regulation [72]. However, it should be noted that our sample excluded individuals with major psychiatric disorders, thereby reducing the range of psychoaffective symptoms severity. Chai et al. [17] proposed a model of temporal progression where sleep disturbances would precede Aβ accumulation, while anxious-depressive symptoms may appear later in AD progression. A complex interplay between these factors would subsequently lead to AD pathology accumulation, neurodegeneration, cognitive decline and dementia. Despite potential interindividual variability in this temporal sequencing, we can hypothesize that anxious-depressive symptoms mostly emerge as a downstream consequence of greater sleep disturbances.

This study has several strengths, including the comprehensive phenotyping of participants for cognitive and behavioral symptoms, as well as amyloid burden. Additionally, the ecological assessment of sleep architecture at home over multiple nights is a novel approach in the field. To our knowledge, this is the first study to investigate night-to-night changes in sleep architecture in relation to early AD pathology. However, limitations include the limited number of recorded nights (n = 4.5 ± 0.8) and the cross-sectional design preventing causal inference. Longitudinal studies are needed to determine whether Aβ-related sleep disturbances result from or contribute to amyloid accumulation over time. Although the Somno-Art^®^ device performs within the expected range for similar wearable devices, its ability to correctly classify sleep stages is a limitation that requires further replication.

In conclusion, this study highlights the importance of considering both the mean and variability of sleep parameters when investigating the association between sleep quality and amyloid pathology in cognitively unimpaired individuals. Shorter sleep duration and greater variability in REM sleep duration appear as early markers of AD pathology and potential targets for interventions aimed at reducing AD risk and promoting healthy aging.

## Data Availability

All data produced in the present study are available upon reasonable request to the authors.

## Acknowledgements

The authors acknowledge all the participants of the study for their contribution. We are grateful to the PPRS company (Laurie Thiesse, Antoine Viola, René Gebel, Dylan Besson, and Baptiste Planat) for their support in Somno-Art^®^ data management, the Cyceron MRI-PET staff for neuroimaging data acquisition, as well as Aurélia Cognet, Valérie Lefranc and Géraldine Poisnel for their administrative support. This work was performed on a facility of France Life Imaging network (grant ANR-11-INBS-0006).

## Consent statement

All participants gave written informed consent prior to the examinations.

## REFERENCES

[1] Dubois B, Villain N, Schneider L, Fox N, Campbell N, Galasko D, et al. Alzheimer Disease as a Clinical-Biological Construct—An International Working Group Recommendation. JAMA Neurol 2024;81(12):1304–1311. 10.1001/jamaneurol.2024.3770

[2] Wang C, Holtzman DM. Bidirectional relationship between sleep and Alzheimer’s disease: role of amyloid, tau, and other factors. Neuropsychopharmacol 2020;45(1):104–120. 10.1038/s41386-019-0478-5

[3] Peter-Derex L, Yammine P, Bastuji H, Croisile B. Sleep and Alzheimer’s disease. Sleep Medicine Reviews 2015;19:29–38. 10.1016/j.smrv.2014.03.007

[4] Zhang Y, Ren R, Yang L, Zhang H, Shi Y, Okhravi HR, et al. Sleep in Alzheimer’s disease: a systematic review and meta-analysis of polysomnographic findings. Transl Psychiatry 2022;12(1):136. 10.1038/s41398-022-01897-y

[5] André C, Martineau-Dussault MÈ, Daneault V, Blais H, Frenette S, Lorrain D, et al. REM sleep is associated with the volume of the cholinergic basal forebrain in aMCI individuals. Alzheimers Res Ther 2023;15:151. 10.1186/s13195-023-01265-y

[6] André C, Bédard MA, Daneault V, Wickens R, Soucy JP, Lorrain D, et al. REM sleep EEG slowing reflects brain cholinergic denervation in aging and Mild Cognitive Impairment. medRxiv 2025. Online Preprint. 10.1101/2025.04.28.25326545

[7] Ju YES, Lucey BP, Holtzman DM. Sleep and Alzheimer disease pathology – a bidirectional relationship. Nat Rev Neurol 2014;10(2):115–9. 10.1038/nrneurol.2013.269

[8] André C, Champetier P, Rehel S, Kuhn E, Touron E, Ourry V, et al. Rapid Eye Movement Sleep, Neurodegeneration, and Amyloid Deposition in Aging. Ann Neurol 2023;93(5):979–90. 10.1002/ana.26604

[9] Ettore E, Bakardjian H, Solé M, Levy Nogueira M, Habert MO, Gabelle A, et al. Relationships between objectives sleep parameters and brain amyloid load in subjects at risk for Alzheimer’s disease: the INSIGHT-preAD Study. Sleep 2019;42(9):zsz137. 10.1093/sleep/zsz137

[10] Spira AP, Gamaldo AA, An Y, Wu MN, Simonsick EM, Bilgel M, et al. Self-reported sleep and β-amyloid deposition in community-dwelling older adults. JAMA Neurol 2013;70(12):1537–1543. 10.1001/jamaneurol.2013.4258

[11] Winer JR, Mander BA, Kumar S, Reed M, Baker SL, Jagust WJ, et al. Sleep Disturbance Forecasts β-Amyloid Accumulation across Subsequent Years. Current Biology 2020;30(21):4291–4298.e3. 10.1016/j.cub.2020.08.017

[12] Ahmed AFHM, Ahmed NMAEM, Elsamany AAA, Mohammadany HEMM, Ibrahim SAM, Mustafa FAM, et al. The Role of Sleep Disturbances in Alzheimer’s Disease Progression: A Systematic Review. Cureus 2025;17(6): e86450. 10.7759/cureus.86450

[13] Bergamo G, Liguori C. Are sleep disturbances modifiable risk factors for mild cognitive impairment and dementia? A systematic review of large studies. Sleep Breath 2025;29(4):269. 10.1007/s11325-025-03421-0

[14] Bubu OM, Brannick M, Mortimer J, Umasabor-Bubu O, Sebastião YV, Wen Y, et al. Sleep, Cognitive impairment, and Alzheimer’s disease: A Systematic Review and Meta-Analysis. Sleep 2017;40(1):zsw032. 10.1093/sleep/zsw032

[15] Lucey BP. It’s complicated: The relationship between sleep and Alzheimer’s disease in humans. Neurobiology of Disease 2020;144:105031. 10.1016/j.nbd.2020.105031

[16] Benca R, Herring WJ, Khandker R, Qureshi ZP. Burden of Insomnia and Sleep Disturbances and the Impact of Sleep Treatments in Patients with Probable or Possible Alzheimer’s Disease: A Structured Literature Review. J Alzheimers Dis 2022;86(1):83–109. 10.3233/JAD-215324

[17] Chai Y, Shokri-Kojori E, Saykin AJ, Yu M. Anxious–depressive symptoms and sleep disturbances across the Alzheimer disease spectrum. Nat Mental Health 2025;3(6):594–612. 10.1038/s44220-025-00416-4

[18] Yiallourou S, Baril AA, Wiedner C, Misialek JR, Kline CE, Harrison S, et al. Sleep architecture and dementia risk in adults: an analysis of 5 cohorts from the Sleep and Dementia Consortium. Sleep 2025;48(9):zsaf129. 10.1093/sleep/zsaf129

[19] André C, Stankeviciute L, Michaelian JC, Antonsdottir IM, Benca RM, Coulthard EJ, et al. International recommendations for sleep and circadian research in aging and Alzheimer’s disease: A Delphi consensus study. Alzheimers Dement 2025;21(10):e70742. 10.1002/alz.70742

[20] André C, Tomadesso C, de Flores R, Branger P, Rehel S, Mézenge F, et al. Brain and cognitive correlates of sleep fragmentation in elderly subjects with and without cognitive deficits. Alzheimers Dement 2019;11(1):142–150. 10.1016/j.dadm.2018.12.009

[21] Baril AA, Picard C, Labonté A, Sanchez E, Duclos C, Mohammediyan B, et al. Day-to-day sleep variability with Alzheimer’s biomarkers in at-risk elderly. Alzheimers Dement 2024;16(1):e12521. 10.1002/dad2.12521

[22] Diem SJ, Blackwell TL, Stone KL, Yaffe K, Tranah G, Cauley JA, et al. Measures of Sleep–Wake Patterns and Risk of Mild Cognitive Impairment or Dementia in Older Women. Am J Geriatr Psychiatry 2016;24(3):248–258. 10.1016/j.jagp.2015.12.002

[23] Fenton L, Isenberg AL, Aslanyan V, Albrecht D, Contreras JA, Stradford J, et al. Variability in objective sleep is associated with Alzheimer’s pathology and cognition. Brain Commun 2023;5(2):fcad031. 10.1093/braincomms/fcad031

[24] Jouvencel A, Baillet M, Meyer M, Dilharreguy B, Lamare F, Pérès K, et al. Night-to-night variability in sleep and amyloid beta burden in normal aging. Alzheimers Dement 2023;15(3):e12460. 10.1002/dad2.12460

[25] Yiallourou SR, Cribb L, Cavuoto MG, Rowsthorn E, Nicolazzo J, Gibson M, et al. Association of the Sleep Regularity Index With Incident Dementia and Brain Volume. Neurology 2024;102(2):e208029. 10.1212/WNL.0000000000208029

[26] Poisnel G, Arenaza-Urquijo E, Collette F, Klimecki OM, Marchant NL, Wirth M, et al. The Age-Well randomized controlled trial of the Medit-Ageing European project: Effect of meditation or foreign language training on brain and mental health in older adults. Alzheimers Dement 2018;4:714–23. 10.1016/j.trci.2018.10.011

[27] André C, Rehel S, Kuhn E, Landeau B, Moulinet I, Touron E, et al. Association of Sleep-Disordered Breathing With Alzheimer Disease Biomarkers in Community-Dwelling Older Adults: A Secondary Analysis of a Randomized Clinical Trial. JAMA Neurol 2020;77(6):716. 10.1001/jamaneurol.2020.0311

[28] Müller-Gärtner HW, Links JM, Prince JL, Bryan RN, McVeigh E, Leal JP, et al. Measurement of Radiotracer Concentration in Brain Gray Matter Using Positron Emission Tomography: MRI-Based Correction for Partial Volume Effects. J Cereb Blood Flow Metab 1992;12(4):571–583. 10.1038/jcbfm.1992.81

[29] Chauveau L, Gonneaud J, Poisnel G, Landeau B, Garnier-Crussard A, Pitel AL, et al. Cardiovascular risk factors are associated with lower posterior-medial network functional connectivity in older adults. Alzheimers Res Ther 2025;17(1):159. 10.1186/s13195-025-01808-5

[30] Klunk WE, Koeppe RA, Price JC, Benzinger TL, Devous MD Sr, Jagust WJ, et al. The Centiloid Project: Standardizing Quantitative Amyloid Plaque Estimation by PET. Alzheimers Dement 2015;11(1):1. 10.1016/j.jalz.2014.07.003

[31] Navitsky M, Joshi AD, Kennedy I, Klunk WE, Rowe CC, Wong DF, et al. Standardization of amyloid quantitation with florbetapir standardized uptake value ratios to the Centiloid scale. Alzheimers Dement 2018;14(12):1565–1571. 10.1016/j.jalz.2018.06.1353

[32] La Joie R, Ayakta N, Seeley WW, Borys E, Boxer AL, DeCarli C, et al. Multisite study of the relationships between antemortem [11C]PIB-PET Centiloid values and postmortem measures of Alzheimer’s disease neuropathology. Alzheimers Dement 2019;15(2):205–216. 10.1016/j.jalz.2018.09.001

[33] Muzet A, Werner S, Fuchs G, Roth T, Saoud JB, Viola AU, et al. Assessing sleep architecture and continuity measures through the analysis of heart rate and wrist movement recordings in healthy subjects: comparison with results based on polysomnography. Sleep Medicine 2016;21:47–56. 10.1016/j.sleep.2016.01.015

[34] Thiesse L, Staner L, Bourgin P, Roth T, Fuchs G, Kirscher D, et al. Validation of Somno-Art Software, a novel approach of sleep staging, compared with polysomnography in disturbed sleep profiles. Sleep Adv 2022;3(1):zpab019. 10.1093/sleepadvances/zpab019

[35] Buman MP, Hekler EB, Bliwise DL, King AC. Exercise Effects on Night-to-Night Fluctuations in Self-rated Sleep among Older Adults with Sleep Complaints. J Sleep Res 2011;20(1 Pt 1):28–37. 10.1111/j.1365-2869.2010.00866.x

[36] Rowe M, McCrae C, Campbell J, Horne C, Tiegs T, Lehman B, et al. Actigraphy in Older Adults: Comparison of Means and Variability of Three Different Aggregates of Measurement. Behavioral Sleep Medicine 2008;6(2):127–145. 10.1080/15402000801952872

[37] van Hilten JJ, Braat EAM, van der Velde EA, Middelkoop HAM, Kerkhof GA, Kamphuisen HAC. Ambulatory Activity Monitoring During Sleep: An Evaluation of Internight and Intrasubject Variability in Healthy Persons Aged 50-98 Years. Sleep 1993;16(2):146–150. 10.1093/sleep/16.2.146

[38] Buysse DJ, Reynolds CF, Monk TH, Berman SR, Kupfer DJ. The Pittsburgh sleep quality index: A new instrument for psychiatric practice and research. Psychiatry Research 1989;28(2):193–213. 10.1016/0165-1781(89)90047-4

[39] Bastien C, Vallières A, Morin C. Validation of the Insomnia Severity Index as an outcome measure for insomnia research. Sleep Medicine 2001;2(4):297–307. 10.1016/S1389-9457(00)00065-4

[40] McNair DM, Kahn RJ. Self-assessment of cognitive deficits. In: Crook T, Ferris S, Bartus R, editors. Assessment in geriatric psychopharmacology. New Canaan: Mark Powley Associates; 1983: 119–136.

[41] Sheikh JI, Yesavage JA. Geriatric Depression Scale (GDS): Recent evidence and development of a shorter version. Clin Gerontol. 5(1-2), 1986: 165–173. 10.1300/J018v05n01_09

[42] Spielberger CD, Gorsuch RL, Lushene RE. Manual for the State-Trait Anxiety Inventory. Palo Alto, CA: Consulting Psychologists Press; 1970.

[43] Treynor W, Gonzalez R, Nolen-Hoeksema S. Rumination Reconsidered: A Psychometric Analysis. Cognitive Therapy and Research 2003;27(3):247–259. 10.1023/A:1023910315561

[44] Meyer TJ, Miller ML, Metzger RL, Borkovec TD. Development and validation of the penn state worry questionnaire. Behaviour Research and Therapy 1990;28(6):487–495. 10.1016/0005-7967(90)90135-6

[45] Chen CL, Zhang MY, Wang ZL, Deng JH, Bao YP, Shi J, et al. Associations among sleep quality, sleep duration, and Alzheimer’s disease biomarkers: A systematic review and meta-analysis. Alzheimers Dement 2025;21(3):e70096. 10.1002/alz.70096

[46] Insel PS, Mohlenhoff BS, Neylan TC, Krystal AD, Mackin RS. Association of Sleep and β-Amyloid Pathology Among Older Cognitively Unimpaired Adults. JAMA Netw Open 2021;4(7):e2117573. 10.1001/jamanetworkopen.2021.17573

[47] Winer JR, Deters KD, Kennedy G, Jin M, Goldstein-Piekarski A, Poston KL, et al. Association of Short and Long Sleep Duration With Amyloid-β Burden and Cognition in Aging. JAMA Neurol 2021;78(10):1187–96. 10.1001/jamaneurol.2021.2876

[48] Alders P, Kok A, van Zutphen EM, Claassen JAHR, Deeg DJH. The effect of sleep disturbances on the incidence of dementia for varying lag times. J Prev Alzheimers Dis 2025;12(2):100024. 10.1016/j.tjpad.2024.100024

[49] Sabia S, Fayosse A, Dumurgier J, van Hees VT, Paquet C, Sommerlad A, et al. Association of sleep duration in middle and old age with incidence of dementia. Nat Commun 2021;12(1):2289. 10.1038/s41467-021-22354-2

[50] Moon C, Schneider A, Cho YE, Zhang M, Dang H, Vu K. Sleep duration, sleep efficiency, and amyloid β among cognitively healthy later-life adults: a systematic review and meta-analysis. BMC Geriatrics 2024;24(1):408. 10.1186/s12877-024-05010-4

[51] Windred DP, Burns AC, Lane JM, Saxena R, Rutter MK, Cain SW, et al. Sleep regularity is a stronger predictor of mortality risk than sleep duration: A prospective cohort study. Sleep 2024;47(1):zsad253. 10.1093/sleep/zsad253

[52] Mukai Y, Yamanaka A. Functional roles of REM sleep. Neuroscience Research 2023;189:44–53. 10.1016/j.neures.2022.12.009

[53] André C, Martineau-Dussault MÈ, Baril AA, Marchi NA, Daneault V, Lorrain D, et al. Reduced rapid eye movement sleep in late middle-aged and older apolipoprotein E ɛ4 allele carriers. Sleep 2024;47(7):zsae094. 10.1093/sleep/zsae094

[54] Pase MP, Himali JJ, Grima NA, Beiser AS, Satizabal CL, Aparicio HJ, et al. Sleep architecture and the risk of incident dementia in the community. Neurology 2017;89(12):1244–1250. 10.1212/WNL.0000000000004373

[55] Song Y, Blackwell T, Yaffe K, Ancoli-Israel S, Redline S, Stone KL. Relationships Between Sleep Stages and Changes in Cognitive Function in Older Men: The MrOS Sleep Study. Sleep 2015;38(3):411–421. 10.5665/sleep.4500

[56] Jin J, Chen J, Cavaillès C, Yaffe K, Winer J, Stankeviciute L, et al. Association of rapid eye movement sleep latency with multimodal biomarkers of Alzheimer’s disease. Alzheimers Dement 2024;n/a(n/a):e14495. 10.1002/alz.14495

[57] Eser RA, Ehrenberg AJ, Petersen C, Dunlop S, Mejia MB, Suemoto CK, et al. Selective Vulnerability of Brainstem Nuclei in Distinct Tauopathies: A Postmortem Study. J Neuropathol Exp Neurol 2018;77(2):149–161. 10.1093/jnen/nlx113

[58] Lew CH, Petersen C, Neylan TC, Grinberg LT. Tau-driven degeneration of sleep- and wake-regulating neurons in Alzheimer’s disease. Sleep Medicine Reviews 2021;60:101541. 10.1016/j.smrv.2021.101541

[59] Mesulam M, Shaw P, Mash D, Weintraub S. Cholinergic nucleus basalis tauopathy emerges early in the aging-MCI-AD continuum. Ann Neurol 2004;55(6):815–828. 10.1002/ana.20100

[60] Oh J, Eser RA, Ehrenberg AJ, Morales D, Petersen C, Kudlacek J, et al. Profound degeneration of wake-promoting neurons in Alzheimer’s disease. Alzheimers Dement 2019;15(10):1253–1263. 10.1016/j.jalz.2019.06.3916

[61] Stratmann K, Heinsen H, Korf HW, Del Turco D, Ghebremedhin E, Seidel K, et al. Precortical Phase of Alzheimer’s Disease (AD)-Related Tau Cytoskeletal Pathology. Brain Pathol 2016;26(3):371–386. 10.1111/bpa.12289

[62] Van Erum J, Van Dam D, De Deyn PP. Alzheimer’s disease: Neurotransmitters of the sleep-wake cycle. Neurosci Biobehav Rev 2019;105:72–80. 10.1016/j.neubiorev.2019.07.019

[63] Shen Y, Lv Qk, Xie Wy, Gong Sy, Zhuang S, Liu Jy, et al. Circadian disruption and sleep disorders in neurodegeneration. Transl Neurodegener 2023;12(1):8. 10.1186/s40035-023-00340-6

[64] Park SH, Weber F. Neural and Homeostatic Regulation of REM Sleep. Front Psychol 2020;11:1662. 10.3389/fpsyg.2020.01662

[65] Deantoni M, Reyt M, Dourte M, de Haan S, Lesoinne A, Vandewalle G, et al. Circadian rapid eye movement sleep expression is associated with brain microstructural integrity in older adults. Commun Biol 2024;7(1):758. 10.1038/s42003-024-06415-y

[66] Liguori C, Nuccetelli M, Izzi F, Sancesario G, Romigi A, Martorana A, et al. Rapid eye movement sleep disruption and sleep fragmentation are associated with increased orexin-A cerebrospinal-fluid levels in mild cognitive impairment due to Alzheimer’s disease. Neurobiology of Aging 2016;40:120–126. 10.1016/j.neurobiolaging.2016.01.007

[67] Mattsson N, Palmqvist S, Stomrud E, Vogel J, Hansson O. Staging β-Amyloid Pathology With Amyloid Positron Emission Tomography. JAMA Neurol 2019;76(11). 10.1001/jamaneurol.2019.2214

[68] van Hattem T, Verkaar L, Krugliakova E, Adelhöfer N, Zeising M, Drinkenburg WHIM, et al. Targeting Sleep Physiology to Modulate Glymphatic Brain Clearance. Physiology 2025;40(3):0. 10.1152/physiol.00019.2024

[69] Varga AW, Wohlleber ME, Giménez S, Romero S, Alonso JF, Ducca EL, et al. Reduced Slow-Wave Sleep Is Associated with High Cerebrospinal Fluid Aβ42 Levels in Cognitively Normal Elderly. Sleep 2016;39(11):2041–2048. 10.5665/sleep.6240

[70] Lo JC, Groeger JA, Cheng GH, Dijk DJ, Chee MWL. Self-reported sleep duration and cognitive performance in older adults: a systematic review and meta-analysis. Sleep Medicine 2016;17:87–98. 10.1016/j.sleep.2015.08.021

[71] Cavaillès C, Carrière I, Wagner M, Dartigues JF, Berr C, Dauvilliers Y, et al. Trajectories of sleep duration and timing before dementia: a 14-year follow-up study. Age Ageing 2022;51(8):afac186. 10.1093/ageing/afac186

[72] Palmer CA, Alfano CA. Sleep and emotion regulation: An organizing, integrative review. Sleep Medicine Reviews 2017;31:6–16. 10.1016/j.smrv.2015.12.006

