## Supplementary Material for "Night-to-night REM sleep variability: a relevant marker of early amyloid-β deposition"

### Supplementary Methods

#### Performance evaluation of Somno-Art® Software

Somno-Art® Software has been validated against polysomnography (PSG) visual scoring in healthy adults and patients with insomnia, obstructive sleep apnea, and major depressive disorder [1],[2]. In order to evaluate its performance in an elderly population, we conducted comparison analyses between Somno-Art® Software classification and PSG visual scoring performed on recordings obtained on a same night. A convenience sample of 106 recorded nights from the Age-Well randomized controlled trial (at two different time points, each subject having only one night in the dataset) was used. Somno-Art® data were obtained using the Somno-Art® wearable device (Generation 1) and analyzed using Somno-Art® Software v.2.4.0 [2.4.1]. PSG sleep recordings were conducted at home using an ambulatory device (Siesta®, Compumedics, Australia) and were scored in 30-s epochs according to American Academy of Sleep Medicine (AASM) criteria. Exclusion criteria for Somno-Art® recordings included unavailable valid PSG matching Somno-Art® recordings, incorrect use of the device, failure to import data, time-stamping issues, invalid recordings for software analysis (i.e., bad actigraphy and/or heart rate signals), or exclusion from quality control (e.g., bad signal quality).

According to the Sleep Research Society "International Biomarkers Workshop on Wearables in Sleep and Circadian Science" guidelines for the validation of wearables for sleep and circadian outcomes and recommendations for their use [3],[4], recommended statistical analyses (i.e., descriptive statistics, discrepancy analysis, epoch-by-epoch (EBE) analysis) were conducted to evaluate the agreement between Somno-Art® Software and PSG. Discrepancy analysis and EBE analysis were performed according to the standardized analytical framework of Menghini et al. [5] for testing the performance of a sleep tracking technology (https://sri-human-sleep.github.io/sleep-trackers-performance/AnalyticalPipeline_v1.0.0.html). Statistical analyses were performed using R (version 4.4.2). The following sleep parameters were computed: Total Sleep Time (TST, min), Sleep Efficiency (SE, %), Sleep Onset Latency (SOL, min), Duration (min) and Percentage (%, relative to time in bed *minus* SOL) of Wake After Sleep Onset (WASO), Duration (min) and Percentage (%, relative to TST) of Light Sleep (N1+N2), Deep Sleep (N3), and Rapid Eye Movement Sleep (REM, min), REM Sleep Latency (REML, min), and Wake Duration (W, min, from lights-off to lights-on).

For EBE analysis, a temporal alignment was performed between hypnograms from the two devices [3],[5]. Timestamp synchronization between Somno-Art® and PSG recordings was first performed using the electrocardiogram signals from both devices, followed by synchronization on a 30-s epoch level. The period between lights-off and lights-on identified by the PSG was considered as the time window for EBE analysis. The unscored periods from Somno-Art® Software were considered as periods of wakefulness as they were outside the period allocated to sleep.

***Discrepancy analysis.*** Individual-level discrepancy analysis was first performed by subtracting reference-derived measures from device-derived measures (i.e., Somno-Art® – PSG), such that a positive difference indicates that Somno-Art® Software overestimated the observed outcome. Conversely, a negative difference indicates that the device underestimated the observed outcome compared to the reference-derived measures. The individual-level discrepancies were then used to estimate the bias and the 95% limits of agreement (LOAs) of Somno-Art®, adjusting for specific cases of non-compliance with the assumptions (see Menghini et al. [5] for more information). Results of group-level discrepancies are reported in Table S1. Bland-Altman plots (Figure S1) allow visualization of discrepancies, in which the differences between Somno-Art® and PSG-derived measures are plotted against size of measurement (SM, expressed as the range of PSG-derived measures). All sleep parameters considered showed a negative proportional bias, the mean difference between each measure obtained with Somno-Art® Software and PSG decreasing as a function of the SM. Thus, the bias cannot be evaluated as significant or non-significant, as it depends on the SM [4]. Broadly speaking, sleep measurements tend to be overestimated in cases showing lower PSG-derived measures, and underestimated in cases showing higher PSG-derived measures. Notably, the bias tends to approach zero for higher SE values and lower values of SOL, WASO Duration and Percentage, N3 Duration, and W Duration. The 95% LOAs quantify the random variability of the differences around the bias, or the interval within which 95% of the individual differences are expected to lie [4],[6]. N1+N2 sleep and N3 sleep percentages showed homoscedastic differences, with most differences predicted to lie within 19.15% and 17.11%, respectively, around the bias. Positive heteroscedastic differences were observed for all remaining parameters, with wider LOAs observed for higher PSG-derived measures.

**TABLE S1** Group-level discrepancies between Somno-Art® Software and PSG

| Sleep Parameter | Device mean (SD) | Reference mean (SD) | Bias [95% CI] | LOAs [95% CI] |
|---|---|---|---|---|
| TST (min) | 362.73 (68.9) | 351.03 (61.38) | 139.92 – 0.37 x ref $b_0$ = [89.92, 192.3], $b_1$ = [-0.51, -0.22] | bias ± ref x 0.41 [0.3, 0.53] |
| SE (%) | 79.16 (11.58) | 76.79 (11.45) | 51.91 – 0.65 x ref $b_0$ = [38.62, 68.03], $b_1$ = [-0.83, -0.48] | bias ± ref x 0.41 [0.3, 0.53] |
| SOL (min) | 23.61 (24.24) | 20.71 (15.13) | 15.87 – 0.63 x ref $b_0$ = [4.87, 24.18], $b_1$ = [-0.98, -0.23] | bias ± ref x 1.83 [1.69, 2.35] |
| WASO (min) | 72.99 (50.24) | 87.58 (54.55) | 31.19 – 0.52 x ref $b_0$ = [16.47, 46.45], $b_1$ = [-0.73, -0.35] | bias ± ref x 1.15 [1.01, 1.31] |
| WASO (%) | 16.64 (11.03) | 19.59 (11.54) | 8.56 – 0.59 x ref $b_0$ = [4.75, 12.37], $b_1$ = [-0.76, -0.42] | bias ± ref x 1.16 [1, 1.3] |
| N1+N2 (min) | 223.31 (48.76) | 215.17 (51.22) | 119.12 – 0.52 x ref $b_0$ = [83.7, 151.78], $b_1$ = [-0.67, -0.34] | bias ± ref x 0.48 [0.39, 0.57] |
| N1+N2 (%) | 62.35 (11.11) | 61.5 (11.08) | 33.07 – 0.52 x ref $b_0$ = [22.35, 43.78], $b_1$ = [-0.7, -0.35] | bias ± 19.15 [16.32, 22.28] |
| N3 (min) | 67.27 (40.34) | 69.88 (35.83) | 39.33 – 0.6 x ref $b_0$ = [23.07, 55.29], $b_1$ = [-0.82, -0.39] | bias ± ref x 1.78 [1.62, 2.4] |
| N3 (%) | 18.02 (9.51) | 19.97 (9.69) | 10.24 – 0.61 x ref $b_0$ = [6.36, 14.13], $b_1$ = [-0.79, -0.44] | bias ± 17.11 [14.72, 19.71] |
| REM (min) | 72.15 (28.57) | 65.97 (23.36) | 45.47 – 0.6 x ref $b_0$ = [31.56, 57.51], $b_1$ = [-0.78, -0.39] | bias ± ref x 0.89 [0.77, 1.05] |
| REM (%) | 19.63 (6.74) | 18.54 (5.19) | 14.99 – 0.75 x ref $b_0$ = [11.15, 18.58], $b_1$ = [-0.96, -0.54] | bias ± ref x 0.74 [0.64, 0.86] |
| REML (min) | 90.8 (50.91) | 81.67 (44.85) | 66.74 – 0.71 x ref $b_0$ = [39.41, 92.18], $b_1$ = [-1.03, -0.37] | bias ± ref x 1.18 [1.02, 1.38] |
| W Duration (min) | 96.6 (58.1) | 108.3 (57.53) | 48.28 – 0.55 x ref $b_0$ = [26.07, 71.27], $b_1$ = [-0.8, -0.35] | bias ± ref x 0.99 [0.86, 1.13] |

NOTE. Results of group-level discrepancy analysis between Somno-Art® Software (device) and PSG-derived measures (reference). Proportional bias (i.e., significant correlation between differences and PSG-derived measures) was observed for all sleep parameters, modeled as a function of SM, with a linear regression with intercept $b_0$ and slope $b_1$. Since heteroscedasticity and/or deviation from normality was observed for all parameters except N1+N2 sleep and N3 sleep percentages, bias and LOAs were computed on log-transformed data for these parameters, and back-transformed to express LOAs as a function of SM. LOAs' 95% CI are reported for the back-transformed slope coefficient. 95% CI for the bias and LOAs were obtained using bootstrapping (10,000 replicates) for all sleep parameters for which deviation from normality was detected (i.e., all parameters except N1+N2, N3, and REM sleep percentages). For detailed methodology, see Menghini et al. [5].

Abbreviations: %, percentage; CI, confidence intervals; LOAs, limits of agreement; min, minutes; PSG, polysomnography; ref, reference-derived measures; REM, rapid eye movement sleep; REML, rapid eye movement sleep latency; SD, standard deviation; SE, sleep efficiency; SM, size of measurement; SOL, sleep onset latency; TST, total sleep time; W, wake; WASO, wake after sleep onset.

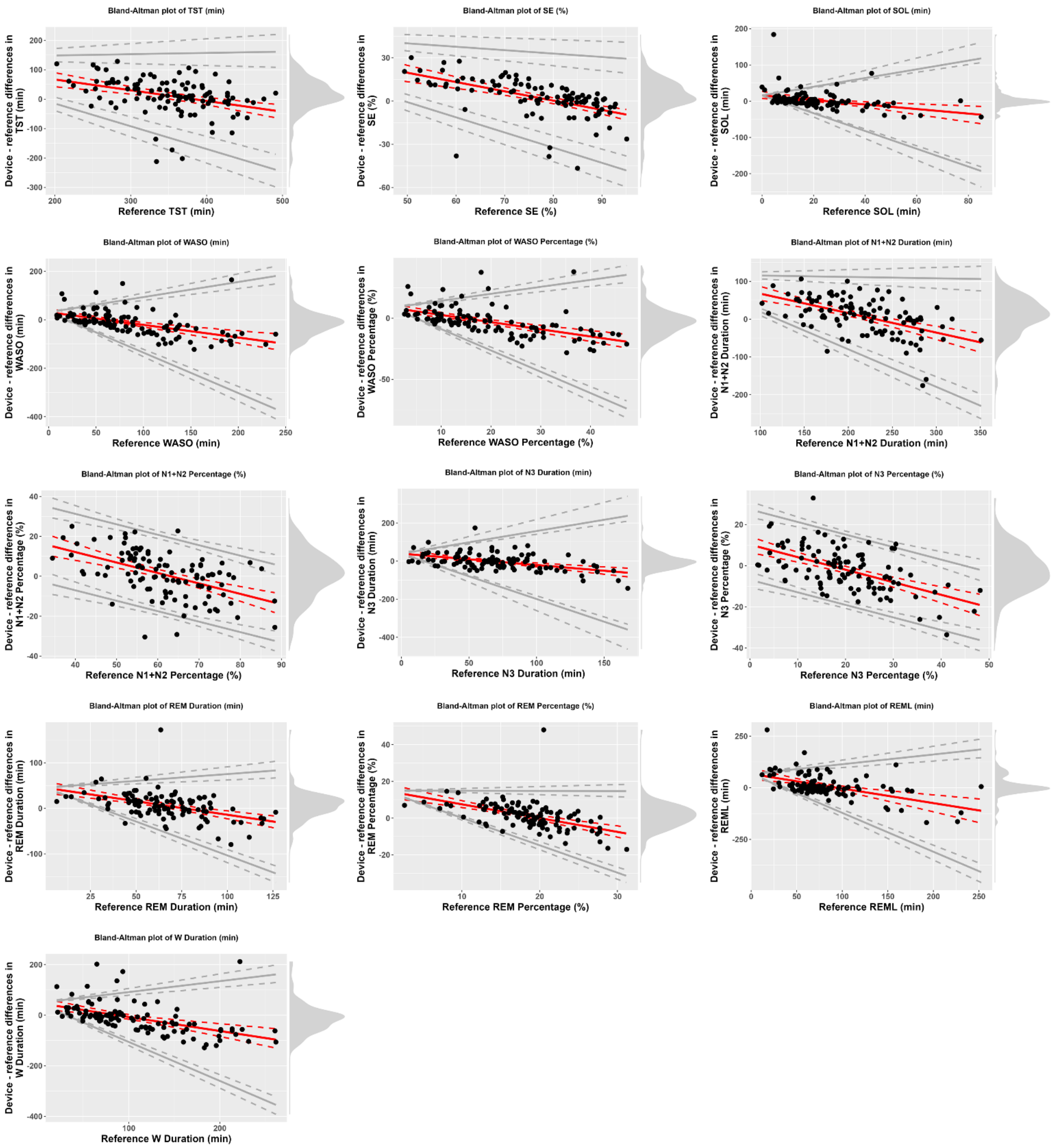

**FIGURE S1** Bland-Altman plots of the sample data. Somno-Art® Software is the device under assessment, and PSG is considered as the reference. Red solid lines represent bias, and gray solid lines represent the 95% LOAs, both with their 95% CIs (dotted lines). Black dots represent individual observations. The density diagram on the right side of each plot illustrates the distribution of the differences. Plots are adjusted for the specific case of compliance with the assumptions for discrepancy analysis: proportional bias, homoscedasticity and normal differences (N1+N2 Percentage, N3 Percentage), proportional bias, heteroscedasticity but normal differences (REM Percentage), and proportional bias, heteroscedasticity and non-normal differences (TST, SE, SOL, WASO, WASO Percentage, N1+N2 Duration, N3 Duration, REM Duration, REML, W Duration). Plots were generated using the analytical pipeline and open-source R code from Menghini et al. [5].

Abbreviations: %, percentage; CI, confidence intervals; LOAs, limits of agreement; min, minutes; PSG, polysomnography; ref, reference-derived measures; REM, rapid eye movement sleep; REML, rapid eye movement sleep latency; SD, standard deviation; SE, sleep efficiency; SOL, sleep onset latency; TST, total sleep time; W, wake; WASO, wake after sleep onset.

***EBE analysis.*** EBE analysis was conducted to assess the accuracy of Somno-Art® Software in both Wake/Sleep classification and W/N1+N2/N3/REM sleep stages classification, compared with PSG visual scoring. Table S2 shows group-level proportional error matrix, providing a representation of EBE analysis by cross-tabulating the agreement and disagreement between Somno-Art® Software and PSG visual scoring for four-stage classification. Table S3 provides EBE accuracy metrics for both Wake/Sleep classification and four-stage classification.

Regarding the Wake/Sleep classification, sensitivity is computed as the proportion of PSG sleep epochs correctly classified by Somno-Art® Software, whereas specificity refers to the ability of Somno-Art® Software to correctly classify PSG wake epochs. Accuracy is the proportion of sleep and wake epochs correctly classified by Somno-Art® over the total number of epochs. For the four-stage classification, sensitivity (i.e., the ability to correctly detect a given sleep stage) is computed as the proportion of epochs classified in a given stage by both Somno-Art® Software and PSG over the total number of epochs classified in that stage by PSG. Specificity, on the other hand, refers to the proportion of epochs classified as any other stage by both methods, divided by the total number of epochs classified as any of the other stages by PSG. Accuracy is the proportion of epochs of a given stage correctly classified by Somno-Art® (i.e., true positive and true negative) over the total number of epochs. The positive predictive value (PPV) is the proportion of epochs that Somno-Art® classifies as a given target stage and that are also classified as that stage by PSG visual scoring, whereas the negative predictive value (NPV) is the proportion of epochs that Somno-Art® does not classify as a target stage and that are not classified as that stage by PSG. Cohen's kappa indicates the proportion of classification agreement for a given sleep stage that is not due to chance. Landis & Kock [7] have proposed a kappa coefficient scale for assessing the agreement between measurements. Coefficients are indicators of "poor" (<0), "slight" (0-0.20), "fair" (0.21-0.40), "moderate" (0.41-0.60), "substantial" (0.61-0.80) and "almost perfect" (>0.81) agreement. Since this metric is sensitive to both the number of categories (i.e., the more categories there are, the lower the kappa) and the prevalence of each condition, the prevalence-adjusted bias-adjusted kappa (PABAK) [8] has also been computed for each stage. PABAK is recommended for evaluating agreement in sleep detection [5]. The prevalence index (PI) is computed as the proportion of epochs that PSG classified as a target stage over the total number of epochs. It quantifies the imbalance in prevalence between the target stage and all non-target stages, higher absolute values reflecting stronger asymmetry. The bias index (BI) is the difference between Somno-Art® and PSG scoring in the proportion of epochs classified as a target stage over the total number of epochs, with values close to zero indicating similar classification proportions. More information about EBE metrics is provided in Menghini et al. [5].

Regarding the Wake/Sleep classification, Somno-Art® Software showed better accuracy and sensitivity than specificity (Table S3). For the four-stage classification, the accuracy coefficients for W, N1+N2, N3, and REM sleep were 81.86%, 64.13%, 85.84%, and 84.08%, respectively (Table S3). The lowest specificity was for N1+N2 sleep (62.47%), along with a lower NPV (67.37%) and PABAK (0.28). As shown in the group-level proportional error matrix (Table S2), confusions between Somno-Art® Software and PSG were mostly due to misclassifications between N1+N2 and N3 sleep, similar to the validation study by Thiesse et al. [2]. Somno-Art® misclassified 30% of W epochs, 45% of N3 epochs and 37% of REM epochs as N1+N2 sleep. This aligns with the lower specificity, NPV and PABAK values obtained for N1+N2 sleep. All other misclassifications ranged from 0.02 to 0.13. According to Landis & Kock [7] kappa coefficient scale, Cohen's kappa coefficients indicated moderate agreement between the two methods for W and fair agreement for all sleep stages. After adjusting for the number and prevalence of categories, however, substantial concordance was observed for W (PABAK: 0.64), N3 (PABAK: 0.72), and REM (PABAK: 0.68) stages, which had the highest absolute PI values.

**TABLE S2** Group-level proportional error matrix between Somno-Art® Software and PSG four-stage classification

| | | **Somno-Art® Software** | | | |
|---|---|---|---|---|---|
| | | **W** | **N1+N2** | **N3** | **REM** |
| **PSG** | **W** | **0.54 (0.19) [0.51, 0.58]** | 0.30 (0.14) [0.28, 0.33] | 0.03 (0.04) [0.02, 0.04] | 0.13 (0.12) [0.10, 0.15] |
| | **N1+N2** | 0.12 (0.12) [0.09, 0.14] | **0.66 (0.12) [0.64, 0.69]** | 0.10 (0.07) [0.09, 0.11] | 0.12 (0.08) [0.10, 0.13] |
| | **N3** | 0.02 (0.08) [0.01, 0.04] | 0.45 (0.23) [0.41, 0.50] | **0.48 (0.26) [0.43, 0.53]** | 0.05 (0.13) [0.02, 0.07] |
| | **REM** | 0.07 (0.15) [0.04, 0.09] | 0.37 (0.21) [0.33, 0.41] | 0.04 (0.09) [0.02, 0.06] | **0.52 (0.25) [0.47, 0.57]** |

NOTE. Results of group-level proportional error matrix, which reports the proportion of Somno-Art®-derived epochs in each condition over the number of PSG-derived epochs in that condition, averaged between subjects, and presented as "mean (SD) [CI]". For detailed methodology, see Menghini et al. [5].

Abbreviations: CI, confidence intervals; PSG, polysomnography; REM, rapid eye movement sleep; SD, standard deviation; W, wake.

**TABLE S3** Group-level EBE metrics for Wake/Sleep classification and four-stage classification

| | **Accuracy (%)** | **Sensitivity (%)** | **Specificity (%)** | **PPV (%)** | **NPV (%)** | **Kappa** | **PABAK** | **BI** | **PI** |
|---|---|---|---|---|---|---|---|---|---|
| ***Wake/Sleep Classification*** | | | | | | | | | |
| Sleep | 81.86 (9.98) [80.01, 83.81] | 90.83 (10.97) [88.87, 93.05] | 54.25 (18.9) [50.68, 57.77] | 86.29 (9.94) [84.37, 88.21] | 66.54 (21.42) [62.52, 70.76] | 0.45 (0.18) [0.42, 0.49] | 0.64 (0.2) [0.6, 0.68] | 0.04 (0.13) [0.01, 0.06] | 0.58 (0.19) [0.55, 0.62] |
| ***Four-stage Classification*** | | | | | | | | | |
| W | 81.86 (9.98) [80.02, 83.77] | 54.25 (18.9) [50.65, 57.89] | 90.83 (10.97) [88.87, 93.03] | 66.54 (21.42) [62.47, 70.64] | 86.29 (9.94) [84.44, 88.22] | 0.45 (0.18) [0.42, 0.49] | 0.64 (0.2) [0.6, 0.68] | -0.04 (0.13) [-0.06, -0.01] | -0.58 (0.19) [-0.62, -0.55] |
| N1+N2 | 64.13 (7.63) [62.7, 65.58] | 66.37 (11.55) [64.28, 68.61] | 62.47 (11.55) [60.27, 64.71] | 61.18 (12.9) [58.75, 63.68] | 67.37 (11.6) [65.21, 69.55] | 0.28 (0.15) [0.25, 0.31] | 0.28 (0.15) [0.26, 0.31] | 0.04 (0.12) [0.02, 0.06] | -0.02 (0.15) [-0.04, 0.01] |
| N3 | 85.84 (5.41) [84.84, 86.91] | 47.81 (25.54) [43.15, 52.66] | 92.92 (4.26) [92.13, 93.71] | 52.39 (27.12) [47.33, 57.6] | 90.73 (6.66) [89.49, 92.03] | 0.39 (0.23) [0.35, 0.43] | 0.72 (0.11) [0.7, 0.74] | -0.02 (0.08) [-0.04, 0] | -0.71 (0.11) [-0.73, -0.69] |
| REM | 84.08 (5.92) [82.97, 85.21] | 51.92 (25.37) [47.17, 56.63] | 89.69 (5.25) [88.7, 90.67] | 44.86 (21.12) [40.88, 48.96] | 91.68 (4.94) [90.77, 92.61] | 0.37 (0.22) [0.33, 0.41] | 0.68 (0.12) [0.66, 0.7] | 0.02 (0.07) [0.01, 0.03] | -0.69 (0.08) [-0.71, -0.68] |

NOTE. Results of group-level basic and advanced EBE metrics for each stage of interest. Individual EBE metrics are averaged between subjects (metricsType = “avg”) and presented as “mean (SD) [CI]”. CI are computed by bootstrap with 10,000 replicates (CI.type=“boot”, boot.type=“basic”). For detailed methodology, see Menghini et al. [5].

Abbreviations: %, percentage; BI, bias index; CI, confidence intervals; EBE, epoch-by-epoch; NPV, negative predictive value; PABAK, prevalence-adjusted bias-adjusted kappa; PI, prevalence index; PPV, positive predictive value; REM, rapid eye movement sleep; SD, standard deviation; W, wake.

In conclusion, when considering all investigated metrics, discrepancy analysis showed a proportional bias for all parameters, the bias magnitude depending on PSG-derived measures. On an EBE basis, Somno-Art® Software showed greater sensitivity than specificity, at 91% and 54%, respectively. These results are consistent with the average reported sensitivity and specificity for wearable devices [3]. Accuracy ranged from 64.13% for N1+N2 sleep to 85.84% for N3 sleep, and agreement between the two methods, as assessed by the PABAK coefficient, ranged from "fair" for N1+N2 to "substantial" for W, N3 and REM stages.

Despite certain limitations in performance, the software helps to address some of the variability that can occur between scorers when performing visual scoring, despite the use of standardized criteria. This is of considerable interest for applications involving multiple sites or repeated measurements such as research [9]. Furthermore, the Somno-Art® device offers the advantage of being easy to use at home, providing sleep architecture measurements over several nights, in the subject's environment.

#### Neuropsychological assessment

To provide robust proxies of cognitive abilities and minimize the issue of multiple statistical testing, composite scores were calculated for each cognitive domain of interest. The raw scores for each test included in a composite score were standardized using the mean and standard deviation of the randomized controlled trial sample (z-scores). The unweighted average of these z-scores then yielded the composite scores. To facilitate the interpretation of longitudinal data, the cognitive composite scores were re-standardized by dividing them by the standard deviation of the baseline data. Note that scores derived from reaction times were reversed so that higher individual scores and therefore higher total composite scores indicate better performance. The tasks used to evaluate each cognitive domain are listed below.

##### *PACC-5 score*

The Preclinical Alzheimer's Cognitive Composite 5 (PACC-5) is a global cognitive composite score sensitive to the detection and tracking of preclinical AD-related decline [10]. It was computed by averaging the standardized scores of the Mattis Dementia Rating Scale-2 (total score) [11], the Coding subtest (raw score) of the Wechsler Adult Intelligence Scale (WAIS)-IV [12], the Category Fluency (number of correct animals produced in 2 minutes) [13], the Logical Memory test – Story B (delayed recall) of the Wechsler Memory Scale (WMS)-IV [14], and the California Verbal Learning Test (CVLT)-II (delayed free recall) [15].

##### *Episodic Memory composite score*

The verbal episodic memory composite score was obtained by averaging the standardized scores of the CVLT-II (3 scores: sum of trials 1-5, immediate free recall, and delayed free recall) [15], and the Logical Memory test – Story B of the WMS-IV (2 scores: immediate and delayed recall) [14].

***Executive Functioning composite score***

The executive functioning composite score was computed as the average of the standardized scores of the Stroop (interference index, i.e., interference response time – color naming response time) [13], the Letter Fluency (number of correct words starting with “p” produced in 2 minutes) [13], the Digit Span backward subtest (raw score) of the WAIS-IV [12], and the Trail Making Test (TMT)-B (response time) [13].

***Attention/speed composite score***

The attention/speed composite score was obtained by averaging the standardized scores of the TMT-A (response time) [13], the Stroop (color naming response time) [13], the Digit Span forward subtest (raw score) of the WAIS-IV [12], and the Coding subtest (raw score) of the WAIS-IV [12].

***Subjective cognitive difficulties***

Subjective cognitive difficulties experienced by participants in their daily activities were assessed with the Cognitive Difficulties Scale [16]. This 39-item self-assessment scale requires participants to rate on a 5-point Likert scale how often they experience particular cognitive difficulties in everyday life (from 0 – ‘Never’ to 4 – ‘Very often’). Five items (1, 20, 27, 30, and 39) were removed from the initial questionnaire because they were either gender-specific (e.g., related to cooking or sewing) or outdated in today's context (e.g., remembering phone numbers). This led to a 34-item reduced score [17]. A 9-item memory score was also calculated to target memory-specific subjective difficulties and consists of items 4, 5, 6, 8, 9, 11, 18, 32, and 35.

#### Psychoaffective evaluation

***Depressive symptoms***

Depressive symptoms were assessed using the 15-item version of the Geriatric Depression Scale (GDS) [18]. This self-reported questionnaire requires subjects to rate how they had felt over the past week on a binary scale. Scores range from 0 to 15, with higher scores indicating more depressive symptoms.

***Trait anxiety***

Trait anxiety was evaluated using Form Y-B of the State-Trait Anxiety Inventory (STAI-B) [19]. This 20-item questionnaire requires participants to rate on a 4-point Likert scale how often they generally experience certain feelings (from 1 – 'Almost never' to 4 – 'Almost always'). Scores range from 20 to 80, with higher scores reflecting the presence of more trait anxiety symptoms.

***Repetitive negative thinking***

*Ruminative brooding*. Self-reported ruminative brooding was assessed with the 5-item brooding subscale of the 22-item Rumination Response Scale (RRS) [20]. Each item is rated on a 4-point Likert scale (from 1 – 'Almost never' to 4 – 'Almost always'). Scores range from 5 to 20, with higher scores indicating greater ruminative brooding.

*Worry.* Self-reported trait worry was assessed using the 16-item Penn State Worry Questionnaire (PSWQ) [21]. Each item is rated on a 5-point Likert scale (from 1 – 'Not at all typical of me' to 5 – 'Very typical of me'). Scores range from 16 to 80, with higher scores indicating greater levels of trait worry.

### Supplementary Results

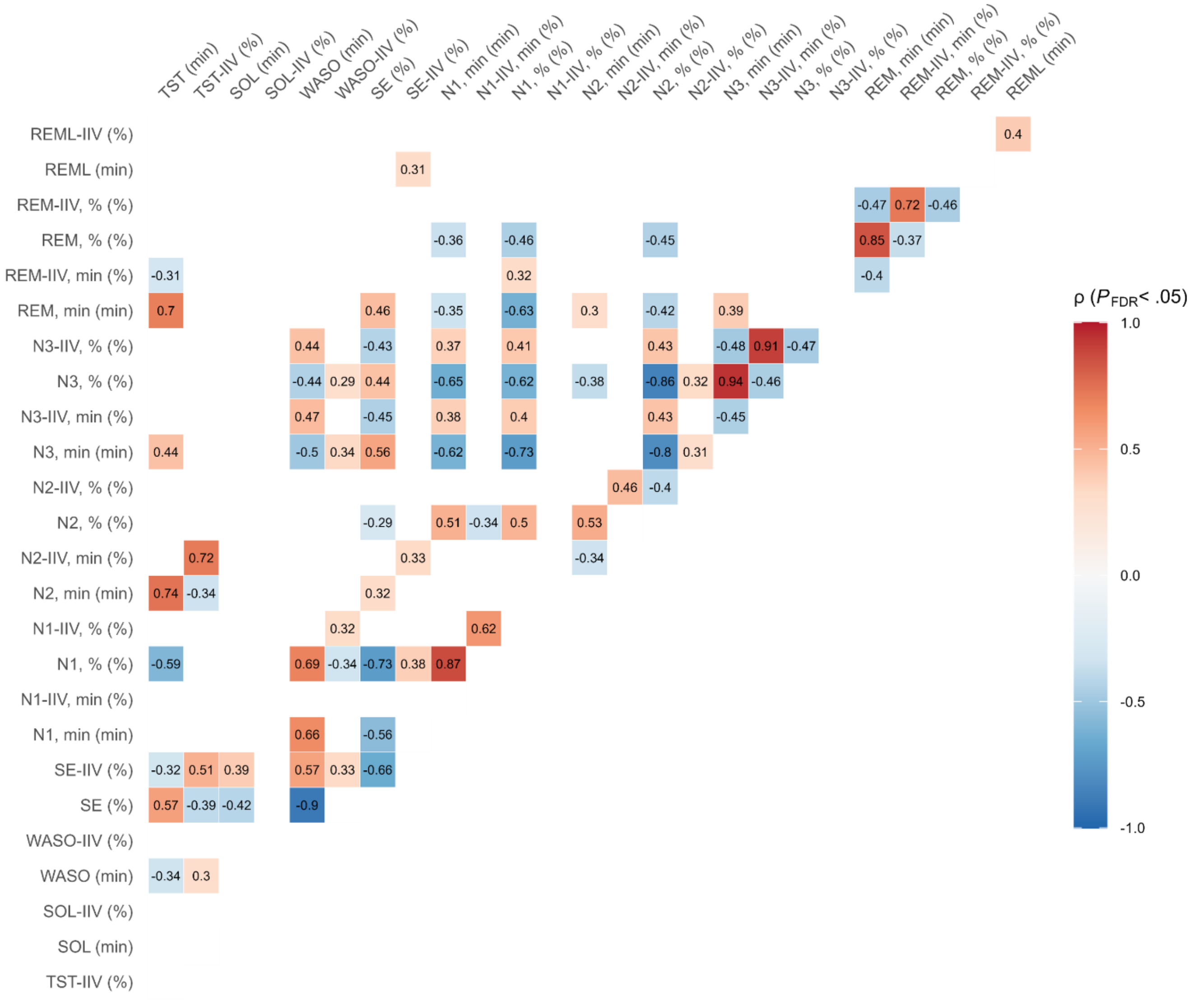

**FIGURE S2** Correlation matrix between sleep metrics. Spearman rho (ρ) is indicated for each significant correlation with a color-coded scale. Results are presented at the $P < .05$ threshold corrected for false discovery rate (FDR).

Abbreviations: %, percentage; IIV, intra-individual variability; min, minutes; REM, rapid eye movement sleep; REML, rapid eye movement sleep latency; SE, sleep efficiency; SOL, sleep onset latency; TST, total sleep time; WASO, wake after sleep onset.

**TABLE S4** Comparison of sleep mean and variability metrics between Aβ- and Aβ+ individuals with robust ANCOVA models

| Sleep parameter | Estimate | Standard Error | *t* | *P* |
|---|---|---|---|---|
| ***Mean Sleep Metrics*** | | | | |
| SOL (min) | -2.0542 | 2.1726 | -0.95 | .348 |
| WASO (min) | -0.4255 | 7.6291 | -0.06 | .956 |
| SE (%) | -0.3180 | 1.6422 | -0.19 | .847 |
| REML (min) | -8.5384 | 9.7389 | -0.88 | .384 |
| ***Intra-Individual Variability (IIV, %) Sleep Metrics*** | | | | |
| TST (min) | 1.9579 | 1.5627 | 1.25 | .214 |
| SOL (min) | 1.5059 | 7.8777 | 0.19 | .849 |
| WASO (min) | -3.7581 | 4.0065 | -0.94 | .351 |
| SE (%) | -0.0421 | 0.8889 | -0.05 | .962 |
| N1 (%) | -3.4628 | 1.8430 | -1.88 | .064 |
| N2 (%) | 1.5247 | 1.4761 | 1.03 | .305 |
| N3 (min) | 1.7409 | 6.2770 | 0.28 | .782 |
| N3 (%) | 0.1608 | 5.6345 | 0.03 | .977 |
| REM (min) | 7.8739 | 2.9209 | 2.70 | **.009** |
| REML (min) | 2.6579 | 7.4937 | 0.35 | .724 |

NOTE. Results of robust ANCOVAs (using *lmrob* function) performed for models that did not meet the conditions for ANCOVA, with each sleep parameter as the dependent variable, Aβ status as the between-participants factor, and age, sex, education, and AHI as covariates. Results in bold indicate significant differences at the *P* <.05 level.

Abbreviations: %, percentage; Aβ, amyloid β; AHI, apnea–hypopnea index; IIV, intra-individual variability; min, minutes; REM, rapid eye movement sleep; REML, rapid eye movement sleep latency; SE, sleep efficiency; SOL, sleep onset latency; TST, total sleep time; WASO, wake after sleep onset.

**TABLE S5** Comparisons of Aβ-related sleep mean and variability metrics between Aβ- and Aβ+ individuals after additional adjustment for *APOE* ε4 status

| Sleep parameter | Df | Aβ- adjusted mean [95% CI] | Aβ+ adjusted mean [95% CI] | F | *P* | $\eta_p^2$ |
|---|---|---|---|---|---|---|
| TST (min) | 1 | 404.22 [389.17, 419.26] | 373.62 [352.22, 395.01] | 5.91 | **.018** | 0.079 |
| REM-IIV, min (%) | 1 | 21.34 [17.88, 24.80] | 28.29 [23.37, 33.21] | 5.77 | **.019** | 0.077 |
| REM-IIV, % (%) | 1 | 18.86 [15.62, 22.09] | 24.25 [19.64, 28.85] | 3.96 | .050 | 0.054 |

NOTE. Results of type II ANCOVAs performed with each Aβ-related sleep parameter (i.e., TST, REM-IIV, min, and REM-IIV, %) as the dependent variable, Aβ status as the between-participants factor, and age, sex, education, AHI, and *APOE* ε4 status as covariates. Results in bold indicate significant differences at the $P < .05$ level.

Abbreviations: %, percentage; Aβ, amyloid β; AHI, apnea–hypopnea index; *APOE* ε4, ε4 allele of the Apolipoprotein E; CI, confidence interval; Df, degrees of freedom; $\eta_p^2$, partial eta squared; IIV, intra-individual variability; min, minutes; REM, rapid eye movement sleep; TST, total sleep time.

**TABLE S6** Comparisons of Aβ-related sleep mean and variability metrics between Aβ- and Aβ+ individuals after additional adjustment for the number of recorded nights

| Sleep parameter | Df | Aβ- adjusted mean [95% CI] | Aβ+ adjusted mean [95% CI] | F | *P* | $\eta_p^2$ |
|---|---|---|---|---|---|---|
| TST (min) | 1 | 396.81 [383.38, 410.25] | 369.48 [348.02, 390.95] | 4.47 | **.038** | 0.061 |
| REM-IIV, min (%) | 1 | 22.42 [19.42, 25.42] | 28.56 [23.76, 33.36] | 4.51 | **.037** | 0.061 |
| REM-IIV, % (%) | 1 | 19.16 [16.46, 21.87] | 23.71 [19.38, 28.03] | 3.04 | .085 | 0.042 |

NOTE. Results of type II ANCOVAs performed with each Aβ-related sleep parameter (i.e., TST, REM-IIV, min, and REM-IIV, %) as the dependent variable, separately, Aβ status as the between-participants factor, and age, sex, education, AHI, and the number of recorded nights as covariates. Results in bold indicate significant differences at the $P < .05$ level.

Abbreviations: %, percentage; Aβ, amyloid β; AHI, apnea–hypopnea index; CI, confidence interval; Df, degrees of freedom; $\eta_p^2$, partial eta squared; IIV, intra-individual variability; min, minutes; REM, rapid eye movement sleep; TST, total sleep time.

**TABLE S7** Comparison of variability in REM sleep duration (REM-IIV, min) between Aβ- and Aβ+ individuals after additional adjustment for *APOE* ε4 status with a robust ANCOVA model

| Sleep parameter | Estimate | Standard Error | *t* | *P* |
|---|---|---|---|---|
| REM-IIV, min (%) | 8.2739 | 2.9801 | 2.78 | **.007** |

NOTE. Results of the robust ANCOVA (using *lmrob* function) performed with variability in REM sleep duration (REM-IIV, min) as the dependent variable, Aβ status as the between-participants factor, and age, sex, education, AHI, and *APOE* ε4 status as covariates, as the model did not meet the conditions for ANCOVA. The result in bold indicates a significant difference at the $P < .05$ level.

Abbreviations: %, percentage; Aβ, amyloid β; AHI, apnea–hypopnea index; *APOE* ε4, ε4 allele of the Apolipoprotein E; IIV, intra-individual variability; min, minutes; REM, rapid eye movement sleep.

**TABLE S8** Results of sensitivity voxel-wise multiple regressions analyses between Aβ-related sleep metrics and amyloid deposition after additional adjustment for *APOE* ε4 status

| Sleep parameter | Cor | Brain regions | Nb of voxels | MNI coordinates x | y | z | t-value | *P* FWE-corrected |
|---|---|---|---|---|---|---|---|---|
| TST (min) | - | R cuneus, occipital sup | 382 | 14 | -88 | 20 | 4.35 | |
| | | L parietal inf, postcentral, supramarginal | 344 | -51 | -26 | 54 | 3.97 | |
| | | L parietal sup/inf, occipital sup/mid, precuneus, cuneus, postcentral | 1,445 | -22 | -52 | 58 | 3.55 | |
| | | R precentral, frontal mid | 102 | 40 | -3 | 46 | 3.51 | |
| | | L temporal mid/inf | 523 | -51 | -58 | -2 | 3.40 | |
| | | R temporal inf/mid, fusiform | 473 | 50 | -56 | -15 | 3.38 | |
| | | R temporal sup | 117 | 68 | -14 | 6 | 3.38 | |
| | | L occipital mid | 109 | -32 | -80 | 16 | 3.29 | |
| | | R temporal pole sup/mid | 166 | 38 | 18 | -28 | 3.25 | |
| | | R occipital mid/sup | 144 | 33 | -75 | 26 | 3.17 | |
| REM-IIV, min (%) | + | RL precuneus, cuneus, temporal sup/mid/inf, temporal pole sup/mid, cingulate post/mid, parahippocampal, parietal sup/inf, angular, supramarginal, occipital sup/mid/inf, calcarine, fusiform, lingual, postcentral, paracentral lobule, amygdala, frontal med orb, OFC med, rectus, ACC sub | 56,787 | -50 | 6 | -40 | 4.90 | <.001 |
| | | R frontal inf oper/tri, frontal inf orb, frontal sup/mid, OFC ant/post, insula, rolandic oper, precentral, hippocampus, ACC pre | | | | | | |
| | | R frontal sup/mid/sup med/inf oper, supp motor area, precentral | 5,389 | 18 | 14 | 62 | 3.95 | .019 |
| | | L insula, frontal inf tri | 335 | -28 | 18 | 12 | 3.66 | |
| | | L putamen, caudate, pallidum, vent str, olfactory, OFC post/med, rectus | 766 | -24 | 16 | 2 | 3.63 | |

| | | | | | | | |
|---|---|---|---|---|---|---|---|
| | | L frontal sup/sup med/mid, supp motor area, precentral | 2,275 | -14 | 10 | 69 | 3.58 |
| | | L frontal sup/mid | 286 | -26 | 54 | 6 | 3.27 |
| | | L frontal sup med/sup | 115 | -9 | 63 | 26 | 3.14 |
| | | R putamen, caudate, vent str, pallidum | 341 | 16 | 18 | -4 | 3.02 |
| | | R frontal sup/sup med | 258 | 20 | 62 | 9 | 2.97 |
| REM-IIV, % (%) | + | R temporal pole sup/mid, parahippocampal, amygdala, OFC post | 1,247 | 30 | 14 | -33 | 4.81 |
| | | L temporal pole sup/mid, parahippocampal, amygdala, temporal mid/inf, fusiform | 1,233 | -44 | 20 | -21 | 3.68 |
| | | L calcarine, lingual, precuneus | 586 | -20 | -54 | 3 | 3.65 |
| | | RL precuneus, cuneus<br>R parietal sup | 749 | 16 | -75 | 52 | 3.64 |
| | | L precuneus, cuneus, parietal sup, occipital sup | 572 | -16 | -76 | 44 | 3.38 |
| | | L postcentral, parietal sup, precuneus, paracentral lobule | 367 | -22 | -45 | 70 | 3.37 |
| | | R precuneus | 145 | 21 | -58 | 27 | 3.29 |
| | | R OFC post/med, frontal inf orb, insula | 114 | 21 | 14 | -27 | 3.21 |
| | | R frontal sup/sup med, supp motor area | 452 | 16 | 26 | 62 | 3.21 |
| | | R occipital mid/sup | 224 | 30 | -75 | 26 | 3.18 |
| | | R temporal mid/sup, | 221 | 70 | -30 | -6 | 3.17 |
| | | L angular, occipital mid | 157 | -42 | -72 | 30 | 2.93 |

NOTE. Results of voxel-wise multiple regressions analyses performed between each Aβ-related sleep metric (i.e., TST, REM-IIV, min, and REM-IIV, %), separately, and Aβ load, controlling for age, sex, education, AHI, and *APOE* ε4 status. Results are presented at the $P < .005$ (uncorrected) threshold, with a minimal cluster size of k = 100 voxels. *P*-values after FWE cluster-level correction are indicated when significant. The first region mentioned corresponds to the statistical peak, and the other regions listed compose the rest of each cluster. T-values and coordinates are indicated for the peak of each cluster.

Abbreviations: %, percentage; ; -, negative correlation; +, positive correlation; Aβ, amyloid β; ACC, anterior cingulate cortex; AHI, apnea–hypopnea index; ant, anterior; *APOE* ε4, ε4 allele of the Apolipoprotein E; Cor, correlation; FWE, family-wise error; inf, inferior; IIV, intra-individual variability; L, left; med, medial; mid, middle; min, minutes; MNI, Montreal Neurological Institute; Nb, number; OFC, orbitofrontal cortex; oper, operculum; orb, orbital; post, posterior; R, right; REM, rapid eye movement sleep; sub, subgenual; sup, superior; supp motor area, supplementary motor area; tri, triangular part; TST, total sleep time; vent str, ventral striatum.

**TABLE S9** Results of sensitivity voxel-wise multiple regressions analyses between Aβ-related sleep metrics and amyloid deposition after additional adjustment for the number of recorded nights

| Sleep parameter | Cor | Brain regions | Nb of voxels | MNI coordinates | | | t-value | $P_{\text{FWE-corrected}}$ |
|---|---|---|---|---|---|---|---|---|
| | | | | x | y | z | | |
| TST (min) | - | R cuneus, occipital sup | 357 | 14 | -88 | 20 | 4.39 | |
| | | L postcentral, parietal inf | 106 | -51 | -26 | 54 | 3.55 | |
| | | L parietal sup/inf, postcentral | 174 | -24 | -51 | 58 | 3.31 | |
| | | L temporal mid/inf, occipital mid | 314 | -50 | -58 | -2 | 3.22 | |
| | | R temporal inf | 257 | 50 | -54 | 15 | 3.19 | |
| | | R occipital mid | 108 | 32 | -75 | 26 | 3.09 | |
| | | L parietal sup/inf, occipital sup/mid, precuneus | 240 | -27 | -72 | 40 | 2.92 | |
| REM-IIV, min (%) | + | L temporal inf/mid/sup, temporal pole mid/sup, parahippocampal, fusiform, amygdala | 3,987 | -50 | 6 | -40 | 4.67 | |
| | | RL temporal mid/sup/inf, precuneus, cuneus, cingulate post/mid, parietal sup/inf, angular, supramarginal, occipital sup/mid/inf, calcarine, fusiform, lingual, paracentral lobule | 32,699 | 70 | -32 | -4 | 4.49 | <.001 |
| | | R temporal pole sup/mid, parahippocampal, amygdala, hippocampus | | | | | | |
| | | L postcentral | | | | | | |
| | | R postcentral, precentral, rolandic oper, supramarginal | 795 | 66 | -9 | 24 | 3.60 | |
| | | R frontal sup/mid/sup med, supp motor area, precentral | 1,931 | 40 | -2 | 48 | 3.57 | |
| | | L insula, frontal inf tri | 184 | -28 | 18 | 12 | 3.41 | |

| | | | | | | | |
|---|---|---|---|---|---|---|---|
| | | L putamen, pallidum | 192 | -24 | 16 | 2 | 3.30 |
| | | L frontal sup/sup med, supp motor area | 484 | -15 | 10 | 64 | 3.20 |
| | | R frontal inf oper/tri, precentral, rolandic oper | 307 | 50 | 8 | 12 | 3.17 |
| | | L frontal sup/mid | 144 | -26 | 52 | 4 | 3.13 |
| | | RL rectus<br>L OFC med, frontal med orb, ACC sub | 328 | -4 | 39 | -20 | 2.98 |
| REM-IIV, % (%) | + | R temporal pole sup/mid, parahippocampal | 838 | 28 | 12 | -33 | 4.35 |
| | | L calcarine, lingual, precuneus | 481 | -21 | -54 | 2 | 3.94 |
| | | L temporal pole sup/mid, parahippocampal, temporal inf, fusiform | 831 | -27 | 8 | -38 | 3.67 |
| | | R occipital mid/sup | 204 | 28 | -78 | 26 | 3.37 |
| | | R parietal sup, precuneus, cuneus | 166 | 16 | -75 | 52 | 3.12 |

NOTE. Results of voxel-wise multiple regressions analyses performed between each Aβ-related sleep metric (i.e., TST, REM-IIV, min, and REM-IIV, %), separately, and Aβ load, controlling for age, sex, education, AHI, and the number of recorded nights. Results are presented at the $P < .005$ (uncorrected) threshold, with a minimal cluster size of k = 100 voxels. *P*-values after FWE cluster-level correction are indicated when significant. The first region mentioned corresponds to the statistical peak, and the other regions listed compose the rest of each cluster. T-values and coordinates are indicated for the peak of each cluster.

Abbreviations: %, percentage; -, negative correlation; +, positive correlation; Aβ, amyloid β; ACC, anterior cingulate cortex; AHI, apnea–hypopnea index; Cor, correlation; FWE, family-wise error; inf, inferior; IIV, intra-individual variability; L, left; med, medial; mid, middle; min, minutes; MNI, Montreal Neurological Institute; Nb, number; OFC, orbitofrontal cortex; oper, operculum; orb, orbital; post, posterior; R, right; REM, rapid eye movement sleep; sub, subgenual; sup, superior; supp motor area, supplementary motor area; tri, triangular part; TST, total sleep time.

**TABLE S10** Associations between Aβ-related sleep metrics and cognitive and psychoaffective outcomes with robust linear models

| Predictor | Outcome | Estimate | Standard Error | *t* | *P* |
|---|---|---|---|---|---|
| ***Cognitive Outcomes*** | | | | | |
| TST (min) | PACC-5 | -0.0006 | 0.0015 | -0.36 | .720 |
| | Episodic memory composite score | -0.0022 | 0.0025 | -0.89 | .375 |
| REM-IIV, min (%) | PACC-5 | -0.0008 | 0.0054 | -0.16 | .876 |
| | Episodic memory composite score | -0.0007 | 0.0096 | -0.07 | .943 |
| | CDS reduced score | -0.2706 | 0.1677 | -1.61 | .111 |
| REM-IIV, % (%) | PACC-5 | 0.0029 | 0.0066 | 0.44 | .658 |
| | Episodic memory composite score | 0.0035 | 0.0103 | 0.34 | .737 |
| | CDS reduced score | -0.2158 | 0.1832 | -1.18 | .243 |
| ***Psychoaffective Outcomes*** | | | | | |
| TST (min) | GDS score | -0.0024 | 0.0030 | -0.80 | .425 |
| | RRS brooding score | -0.0041 | 0.0053 | -0.77 | .441 |
| | PSWQ score | -0.0175 | 0.0288 | -0.61 | .544 |
| REM-IIV, min (%) | GDS score | 0.0282 | 0.0187 | 1.51 | .135 |
| | PSWQ score | 0.0864 | 0.0931 | 0.93 | .357 |
| REM-IIV, % (%) | RRS brooding score | 0.0378 | 0.0263 | 1.44 | .155 |
| | PSWQ score | 0.0128 | 0.1247 | 0.10 | .919 |

NOTE. Results of robust multiple linear regressions (using *lmrob* function) performed for models that did not meet the conditions for multiple linear regression, with each Aβ-related sleep metric (i.e., TST, REM-IIV, min, and REM-IIV, %) as the predictor, separately, and cognitive and psychoaffective scores as outcomes, controlling for age, sex, education, and AHI.

Abbreviations: %, percentage; Aβ, amyloid β; AHI, apnea–hypopnea index; CDS, Cognitive Difficulties Scale; GDS, Geriatric Depression Scale; IIV, intra-individual variability; min, minutes; PACC-5, Preclinical Alzheimer's Cognitive Composite 5; PSWQ, Penn State Worry Questionnaire; REM, rapid eye movement sleep; RRS, Rumination Response Scale; TST, total sleep time.

**TABLE S11** Association between Total Sleep Time and CDS memory score after additional adjustment for *APOE* ε4 status

| Predictor | Outcome | Estimate [95% CI] | Standard Error | *t* | *P* |
|---|---|---|---|---|---|
| TST (min) | CDS memory score | 0.025 [0.003, 0.047] | 0.011 | 2.27 | **.026** |

NOTE. Results of the multiple linear regression performed with TST as the predictor, and CDS memory score as the outcome, controlling for age, sex, education, AHI, and *APOE* ε4 status. The result in bold indicates a significant difference at the $P < .05$ level.

Abbreviations: %, percentage; AHI, apnea–hypopnea index; *APOE* ε4, ε4 allele of the Apolipoprotein E; CDS, Cognitive Difficulties Scale; CI, confidence interval; min, minutes; TST, total sleep time.

**TABLE S12** Associations between Total Sleep Time and CDS memory score after additional adjustment for the number of recorded nights

| Predictor | Outcome | Estimate [95% CI] | Standard Error | *t* | *P* |
|---|---|---|---|---|---|
| TST (min) | CDS memory score | 0.024 [0.002, 0.045] | 0.011 | 2.19 | **.032** |

NOTE. Results of the multiple linear regression performed with TST as the predictor, and CDS memory score as the outcome, controlling for age, sex, education, AHI, and the number or recorded nights. The result in bold indicates a significant difference at the $P < .05$ level.

Abbreviations: %, percentage; AHI, apnea–hypopnea index; CDS, Cognitive Difficulties Scale; CI, confidence interval; min, minutes; TST, total sleep time.

### Appendix

#### Appendix A. The Medit-Ageing Research Group

| Name | Affiliation/Location | Role | Contribution |
|---|---|---|---|
| Eider M. Arenaza-Urquijo, PhD | Institut National de la Santé et de la Recherche Médicale, Caen, France | Postdoctoral researcher | Study design; Acquisition, analysis, or interpretation of data |
| Florence Allais, BA | EUCLID/F-CRIN Clinical Trials Platform, Bordeaux, France | Data manager | Data management |
| Julien Asselineau, PhD | EUCLID/F-CRIN Clinical Trials Platform, Bordeaux, France | Statistician | Analysis of data |
| Sebastian Baez Lugo, MSc | University of Geneva, Geneva, Switzerland | PhD student | Acquisition, analysis, or interpretation of data |
| Alexandre Bejanin, PhD | Institut National de la Santé et de la Recherche Médicale, Caen, France | Postdoctoral researcher | Acquisition, analysis, or interpretation of data |
| Anne Chocat, MD | Institut National de la Santé et de la Recherche Médicale, Caen, France | Physician | Inclusion of participants |
| Fabienne Collette, PhD | University of Liege, Liege, Belgium | Group leader | Obtained funding; Study design |
| Sophie Dautricourt, MD, MSc | Institut National de la Santé et de la Recherche Médicale, Caen, France | PhD student | Acquisition, analysis, or interpretation of data |
| Eglantine Ferrand-Devouge, MD, MSc | Institut National de la Santé et de la Recherche Médicale, Caen, France | Physician | Inclusion of participants |
| Robin De Flores, PhD | Institut National de la Santé et de la Recherche Médicale, Caen, France | Postdoctoral Researcher | Acquisition, analysis, or interpretation of data; Administrative, technical, or material support |
| Hélène Esperou, MD | Institut National de la Santé et de la Recherche Médicale, Paris, France | Group leader | Sponsor |
| Eric Frison, MD | EUCLID/F-CRIN Clinical Trials Platform, Bordeaux, France | Methodologist | Study design; Interpretation of data |
| Julie Gonneaud, PhD | Institut National de la Santé et de la Recherche Médicale, Caen, France | Postdoctoral Researcher | Acquisition, analysis, or interpretation of data; Administrative, technical, or material support |
| Olga Klimecki, PhD | University of Geneva, Geneva, Switzerland | Neurologist | Obtained funding; Study design |

| | | | |
|---|---|---|---|
| Valérie Lefranc, BA | Institut National de la Santé et de la Recherche Médicale, Caen, France | Clinical research assistant | Acquisition, analysis, or interpretation of data; Administrative, technical, or material support |
| Antoine Lutz, PhD | Institut National de la Santé et de la Recherche Médicale, Lyon, France | Group leader | Obtained funding; Study design |
| Natalie Marchant, PhD | University College London, United Kingdom | Group leader | Obtained funding; Study design |
| Jose-Luis Molinuevo, MD, PhD | Institut d'Investigacions Biomèdiques August Pi i Sunyer, Barcelona, Spain | Group leader | Obtained funding; Study design |
| Léo Paly, MSc | Institut National de la Santé et de la Recherche Médicale, Caen, France | Neuropsychol ogist | Acquisition, analysis, or interpretation of data |
| Géraldine Poisnel, PhD | Institut National de la Santé et de la Recherche Médicale, Caen, France | Research engineer /Researcher | Obtained funding; Study design; Acquisition, analysis, or interpretation of data; Administrative, technical, or material support |
| Florence Requier, MSc | University of Liege, Liege, Belgium | PhD student | Acquisition, analysis, or interpretation of data |
| Eric Salmon, MD, PhD | University of Liege, Liege, Belgium | Group leader | Obtained funding; Study design |
| Siya Sherif, PhD | Institut National de la Santé et de la Recherche Médicale, Caen, France | Research engineer | Acquisition, analysis, or interpretation of data; Administrative, technical, or material support |
| Edelweiss Touron, PhD | Institut National de la Santé et de la Recherche Médicale, Caen, France | PhD student | Acquisition, analysis, or interpretation of data |
| Matthieu Vanhoutte, PhD | Institut National de la Santé et de la Recherche Médicale, Caen, France | Postdoctoral researcher | Acquisition, analysis, or interpretation of data; Administrative, technical, or material support |
| Patrik Vuilleumier, MD | University of Geneva, Geneva, Switzerland | Group leader | Obtained funding; Study design |
| Miranka Wirth, PhD | Deutsches Zentrum für Neurodegenerative Erkrankungen, Dresden, Germany | Group leader | Study design |

### References

[1] Muzet A, Werner S, Fuchs G, Roth T, Saoud JB, Viola AU, et al. Assessing sleep architecture and continuity measures through the analysis of heart rate and wrist movement recordings in healthy subjects: comparison with results based on polysomnography. Sleep Medicine 2016;21:47-56. https://doi.org/10.1016/j.sleep.2016.01.015

[2] Thiesse L, Staner L, Bourgin P, Roth T, Fuchs G, Kirscher D, et al. Validation of Somno-Art Software, a novel approach of sleep staging, compared with polysomnography in disturbed sleep profiles. Sleep Adv 2022a;3(1):zpab019. https://doi.org/10.1093/sleepadvances/zpab019

[3] Depner CM, Cheng PC, Devine JK, Khosla S, de Zambotti M, Robillard R, et al. Wearable technologies for developing sleep and circadian biomarkers: a summary of workshop discussions. Sleep 2020;43(2):zsz254. https://doi.org/10.1093/sleep/zsz254

[4] de Zambotti M, Goldstein C, Cook J, Menghini L, Altini M, Cheng P, et al. State of the science and recommendations for using wearable technology in sleep and circadian research. Sleep 2024;47(4):zsad325. https://doi.org/10.1093/sleep/zsad325

[5] Menghini L, Cellini N, Goldstone A, Baker FC, de Zambotti M. A standardized framework for testing the performance of sleep-tracking technology: step-by-step guidelines and open-source code. Sleep 2021;44(2):zsaa170. https://doi.org/10.1093/sleep/zsaa170

[6] Bland JM, Altman DG. Statistical methods for assessing agreement between two methods of clinical measurement. Lancet 1986;1(8476):307–310. https://doi.org/10.1016/S0140-6736(86)90837-8

[7] Landis JR, Koch GG. The measurement of observer agreement for categorical data. Biometrics 1977;33(1):159-174. https://doi.org/10.2307/2529310

[8] Byrt T, Bishop J, Carlin JB. Bias, prevalence and kappa. Journal of Clinical Epidemiology 1993;46(5):423–429. https://doi.org/10.1016/0895-4356(93)90018-V

[9] Thiesse L, Staner L, Fuchs G, Kirscher D, Dehouck V, Roth T, et al. Performance of Somno-Art Software compared to polysomnography interscorer variability: A multi-center study. Sleep Medicine 2022b;96:14-19. https://doi.org/10.1016/j.sleep.2022.04.013

[10] Papp KV, Rentz DM, Orlovsky I, Sperling RA, Mormino EC. Optimizing the preclinical Alzheimer's cognitive composite with semantic processing: The PACC5. Alzheimers Dement (N Y). 2017;3(4):668-677. https://doi.org/10.1016/j.trci.2017.10.004

[11] Mattis S. Mental Status Examination for organic mental syndrome in the elderly patient. Geriatr Psychiatry. In: Bellak L, Karasu TB, editors. Geriatric Psychiatry: A Handbook for Psychiatrists and Primary Care Physicians. New York: Grune et Stratton; 1976: 77–121.

[12] Wechsler D. Pearson, Wechsler Adult Intelligence Scale – Fourth Edition (WAIS-IV). Pearson Education, Inc 2008. www.pearsonclinical.co.uk

[13] Godefroy O, GREFEX. Fonctions exécutives et pathologies neurologiques et psychiatriques : évaluation en pratique clinique. Marseille: Solal; 2008.

[14] Wechsler D. Pearson, Wechsler Memory Scale – Fourth Edition (WMS-IV), Pearson Education, Inc 2009. www.pearsonclinical.co.uk

[15] Delis D, Kramer J, Kaplan E, Ober, B. Pearson, California Verbal Learning Test - Second Edition (CVLT-II). Pearson Education, Inc 2000. www.pearsonclinical.co.uk

[16] McNair DM, Kahn RJ. Self-assessment of cognitive deficits. In: Crook T, Ferris S, Bartus R, editors. Assessment in geriatric psychopharmacology. New Canaan: Mark Powley Associates; 1983: 119-136.

[17] Kuhn E, Moulinet I, Perrotin A, La Joie R, Landeau B, Tomadesso C, et al. Cross-sectional and longitudinal characterization of SCD patients recruited from the community versus from a memory clinic: subjective cognitive decline, psychoaffective factors, cognitive

performances, and atrophy progression over time. Alz Res Therapy 2019;11(1):61. https://doi.org/10.1186/s13195-019-0514-z

[18] Sheikh JI, Yesavage JA. Geriatric Depression Scale (GDS): Recent evidence and development of a shorter version. Clin Gerontol. 5(1-2), 1986: 165–173. https://doi.org/10.1300/J018v05n01_09

[19] Spielberger CD, Gorsuch RL, Lushene RE. Manual for the State-Trait Anxiety Inventory. Palo Alto, CA: Consulting Psychologists Press; 1970.

[20] Treynor W, Gonzalez R, Nolen-Hoeksema S. Rumination Reconsidered: A Psychometric Analysis. Cognitive Therapy and Research 2003;27(3):247-259. https://doi.org/10.1023/A:1023910315561

[21] Meyer TJ, Miller ML, Metzger RL, Borkovec TD. Development and validation of the penn state worry questionnaire. Behaviour Research and Therapy 1990;28(6):487-495. https://doi.org/10.1016/0005-7967(90)90135-6
